# Saturation genome editing of DDX3X clarifies pathogenicity of germline and somatic variation

**DOI:** 10.1101/2022.06.10.22276179

**Authors:** E.J. Radford, H.K. Tan, M.H.L. Andersson, J.D Stephenson, E.J. Gardner, H. Ironfield, A.J. Waters, D. Gitterman, S. Lindsay, F. Abascal, I. Martincorena, A. Kolesnik, E. Ng-Cordell, H.V. Firth, K. Baker, J.R.B. Perry, D.J. Adams, S.S. Gerety, M.E. Hurles

## Abstract

Loss-of-function of *DDX3X* is a leading cause of neurodevelopmental disorders (NDD) in females. *DDX3X* is also a somatically mutated cancer driver gene proposed to have tumour promoting and suppressing effects. We performed saturation genome editing of *DDX3X,* testing *in vitro* the functional impact of 12,776 nucleotide variants. We identified 3,432 functionally abnormal variants, in three distinct classes. We trained a machine learning classifier to identify functionally abnormal variants of NDD-relevance. This classifier has at least 97% sensitivity and 99% specificity to detect variants pathogenic for NDD, substantially out-performing *in silico* predictors, and resolving up to 93% of variants of uncertain significance. Moreover, functionally-abnormal variants could account for almost all of the excess nonsynonymous *DDX3X* somatic mutations seen in *DDX3X*- driven cancers. Systematic maps of variant effects generated in experimentally tractable cell types have the potential to transform clinical interpretation of both germline and somatic disease-associated variation.

## Introduction

The disease relevance of the vast majority of variants in disease-associated genes is not known. Predicting the pathogenicity of genetic variants, required for both genetic diagnosis and the identification of cancer driver variants, is challenging. In neurodevelopmental disorders (NDD), this can be particularly difficult, as many phenotypes are non-specific, with hundreds of known disease-associated genes and hundreds yet to be identified^1, 2^. In diagnostic practice, genetic variants are commonly classified according to the American College of Medical Genetics (ACMG/AMP) guidelines, combining information across different types of evidence^3, 4^. Variants with insufficient or conflicting evidence of their effect are classified as variants of uncertain significance (VUS). With increased diagnostic use of genomic sequencing, the number of VUS has grown rapidly^5^. Classification as a VUS precludes diagnosis, provision of accurate recurrence risks for family members, access to targeted treatments, predictive testing, condition-specific support groups, and the psychological benefits of a diagnosis.

Conventional strategies to resolve VUS rely on the accumulation of clinical and population data, use of *in silico* predictors of variant effect, or small-scale retrospective functional studies. Clinical data accumulates too slowly to usefully resolve most VUS in rare developmental disorders. Current approaches to generating informative functional data are too slow and expensive to meet the growing need. *In silico* predictors are widely utilised, but these tools often produce conflicting results and can only modestly influence variant classification under the ACMG/AMP guidelines.

Multiplexed assays of variant effect (MAVEs) that empirically and prospectively assess variant effects at scale have numerous advantages over small-scale retrospective functional studies^5^. The effect of an individual variant is interpreted in the context of the effects of many other possible variants in the same gene. This allows estimation of the assay sensitivity and specificity, and an empirical calibration for weighting of the information within diagnostic pathways^6^. Saturation genome editing (SGE) is a MAVE technique utilising CRISPR/Cas9- stimulated homology directed repair (HDR) to introduce specified genetic variants into an endogenous locus. SGE of essential genes in the near-haploid HAP1 cell line identifies variants which impair the expression or function of that gene through variant depletion in cell culture over time. Importantly, because variants are introduced into the endogenous locus, genomic and regulatory context is preserved, therefore non-coding and synonymous variants which alter splicing or transcriptional control can also be detected^7^. SGE has been demonstrated to effectively resolve most VUS in *BRCA1*^7, 8^, outperforms *in silico* prediction algorithms^9^, and has proven diagnostic utility^8, 10^.

Pathogenic variation in *DDX3X*, an X-linked gene encoding an RNA helicase of the DEAD-box protein family, has been robustly associated with both NDD (germline variation) and cancer (somatic mutations). Heterozygous *DDX3X* loss-of-function variants are one of the most common genetic causes of intellectual disability in females^1, 11^, with an approximately equal split between protein-truncating and missense variants. However, the relatively variable and non-specific presentation can hamper accurate variant interpretation. Protein-truncating variants are not seen in males, presumably because they are lethal^12^. It has been proposed that some *DDX3X* missense variants in males can also cause NDD^13^, however, these are observed much less frequently than in females and the statistical evidence for mutation enrichment in NDD-affected individuals is not conclusive (previously reported p-values for mutation enrichment in females 7.4×10^-96^, in males 1.5×10^-3^)^14^. *DDX3X* has also been identified as a somatic driver gene in several different cancers, with the most statistically compelling evidence of somatic mutation enrichment in medulloblastoma^15–17^. Uncertainty persists, however, as to whether *DDX3X* acts as an oncogene or tumour suppressor, particularly in medulloblastoma^18, 19^.

Several studies have functionally characterised *DDX3X* pathogenic variants observed in individuals with NDDs, using a variety of model systems^11, 15, 20–22^. The largest of these involved quantifying the impact of eight variants on helicase activity *in vitro*^20^. Functional experiments in zebrafish have suggested one missense variant seen in an affected male might result in partial loss-of-function^21^. *In vitro* functional characterisation of one pathogenic missense variant seen in an affected female suggests that it might operate via a toxic protein aggregation mechanism^22^.

*DDX3X* was identified as an essential gene in a genome-wide CRISPR essentiality screen performed in the HAP1 cell line^23, 24^, suggesting that *DDX3X* might be suitable for SGE in HAP1 cells. Here we characterise the functional consequence of over 12,000 *DDX3X* variants, using an improved SGE-based assay, to assess their relevance for NDDs and cancer.

## Results

To improve the sensitivity in the SGE assay described in Findlay *et. al*., we made several refinements. Most notably, we increased the transfection efficiency in HAP1 cells to 40-50% (Extended Data 1A), by i) reducing the size of the transfected plasmids, ii) integrating the large Cas9 expressing construct into the cell prior to SGE and iii) re-optimizing the transfection conditions. High transfection efficiencies should increase the representation of the variants being tested, improving signal-to-noise ratio in our SGE assay.

In a pilot SGE study of *DDX3X*, we observed a strong depletion of all nonsense variants relative to synonymous variants on Day 11 in exon 14, confirming that *DDX3X* is essential in HAP1 cells. It also suggested that our adapted SGE protocol generates data with high signal-to-noise ratio (Extended Data 1B).

To perform SGE on all 17 coding exons of *DDX3X*, we designed two independent experiments per exon, using different sgRNAs and HDR template libraries (Fig.1A), with each experiment performed in triplicate. Each HDR template contained a unique variant to be assessed and 1-3 synonymous variants in the sgRNA PAM region and protospacer (common between all HDR templates in the same library) to prevent Cas9 re-cutting. To characterise the time course of relative variant abundance in culture (Fig.1B), cells were harvested at five timepoints (Day 4, 7, 11, 15 and 21 post-transfection), and targeted sequencing of the relevant exon was performed on DNA extracted at each timepoint.

**Fig. 1.**
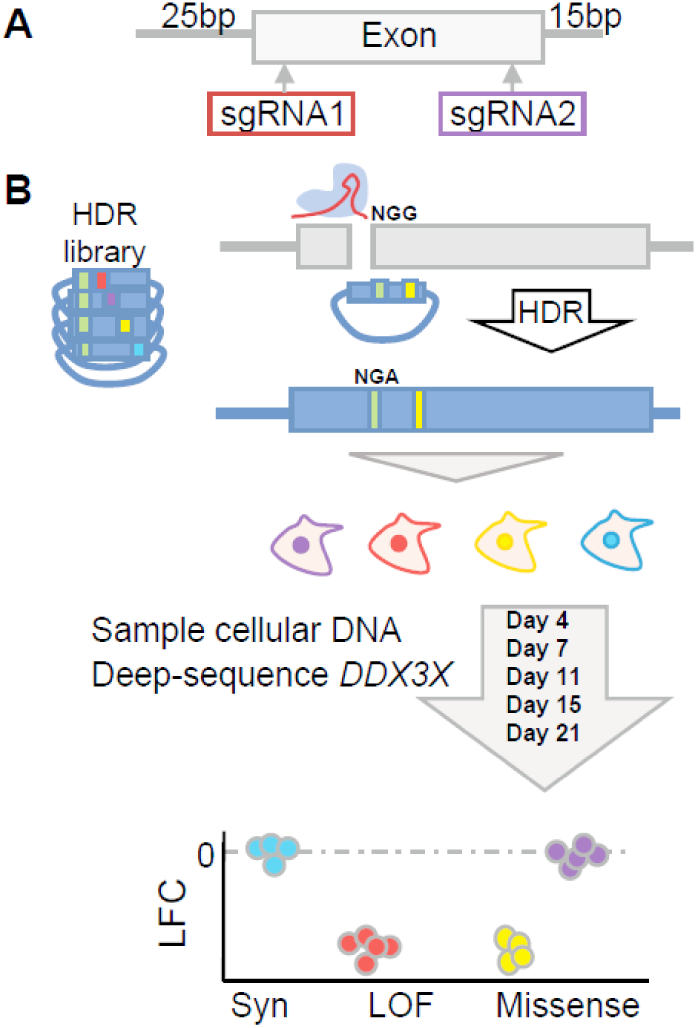
Experimental design. A) For each exon two independent sgRNAs and associated HDR variant libraries are designed at the 5’ and 3’ end. B) The sgRNA, together with the HDR variant library are transfected into *LIG4*-KO Cas9-expressing HAP1 cells. The sgRNA directs the Cas9-mediated double-stranded DNA cut to the target exon. HDR utilises the plasmid library as a template for repair, incorporating a single *DDX3X* variant of interest into the endogenous locus of each cell. Each donor template also carries 1-3 synonymous changes to the NGG PAM site and protospacer, preventing re-cutting. As *DDX3X* is essential, variants which abrogate gene function will cause those cells to die. We sample cells on days 4,7,11,15 and 21 and deep-sequence the genomic DNA to quantify variants’ abundance. We anticipate that functional missense (purple) and synonymous variants (Syn, blue) remain abundant, while loss-of-function variants (LOF, red), and deleterious missense (yellow) variants are depleted from culture.

Collectively, the HDR template libraries contained all possible 6,276 coding single nucleotide variants (SNVs); all 626 deletions of contiguous codons; all 51 insertions and deletions less than 50bp reported in ClinVar, DECIPHER, GnomAD or UK Biobank; and 2,127 non-coding SNVs and 2bp deletions located within 25bp 5’ and 15bp 3’ of the exon boundary. In addition, for each coding SNV, at least one ‘redundant variant’ was designed: a variant resulting in the same amino acid change with different nucleotide changes in that codon, typically a multi-nucleotide variant (3,696 variants). Including the redundant variants allowed us to assess the concordance of the effect of the same amino-acid change generated by two different genetic variants.

Sequencing data were processed to generate counts of each introduced variant at all assayed timepoints, in triplicate, which were subsequently analysed using DESeq2^25^ using Day 4 samples as a baseline for log2 fold-change calculation (LFC) for each variant at each timepoint. To integrate data across all timepoints, DESeq2 was adapted to model the change in LFC across timepoints, generating a single functional score (LFC-trend) for each variant corresponding to the rate of change in relative abundance over time. For each exon, the LFC and LFC-trend scores of variants were normalised against the median of the synonymous and intronic variants (not including canonical splice sites). We observed a high correlation between the two independent sgRNA experiments within each exon for both measures of variant abundance (Pearson’s correlation: LFC-trend = 0.897, LFC: 0.886 to 0.913 for each timepoint) (Extended Data 2A-E). Measures of variant abundance for the same variant across the two sgRNA experiments for each exon were combined using a weighted mean. The resultant combined LFC-trend score (cLFC-trend) is highly correlated with the combined-LFC (cLFC) of variant abundance at Day 15 (Pearson r=0.995, Extended Data 2F). The statistical significance of the cLFC of a variant differing from that of synonymous and intronic variants within the same exon was quantified using the Benjamini-Hochberg (BH) corrected False Discovery Rate (FDR).

We identified 2,337 significantly (Day 15 cLFC FDR < 0.01) SGE-depleted variants and 1,095 significantly (Day 15 cLFC FDR <= 0.01) SGE-enriched variants. Both classes exhibited good concordance between redundant variants: 95% of SGE-depleted variants had a redundant codon that was also classified as SGE-depleted, while 69% of SGE-enriched variants had a redundant codon that was also classified as SGE-enriched, with directional consistencies of 99% and 95% respectively (Extended Data 3). Variants with cLFC FDR > 0.01 on Day 15 were classified as “SGE-unchanged”. We had not anticipated observing a class of SGE-enriched variants, however, missense variants are very significantly overrepresented among SGE-enriched variants, by 1.5-fold (□^2^ p=3.2×10^-36^), suggesting that they are unlikely to represent technical artefacts. Significantly depleted variants were further subdivided into slow-depleting and fast-depleting based on a two-dimensional Gaussian mixture model of Day 7 and Day 15 cLFCs (Fig.2A, B). Day 15 cLFC was used in preference to Day 21 cLFC as a subtle increase in mean variant abundance was observed from Day 15 to Day 21, likely due to increased diploidy within the assayed cell population at this later time point.

**Fig. 2.**
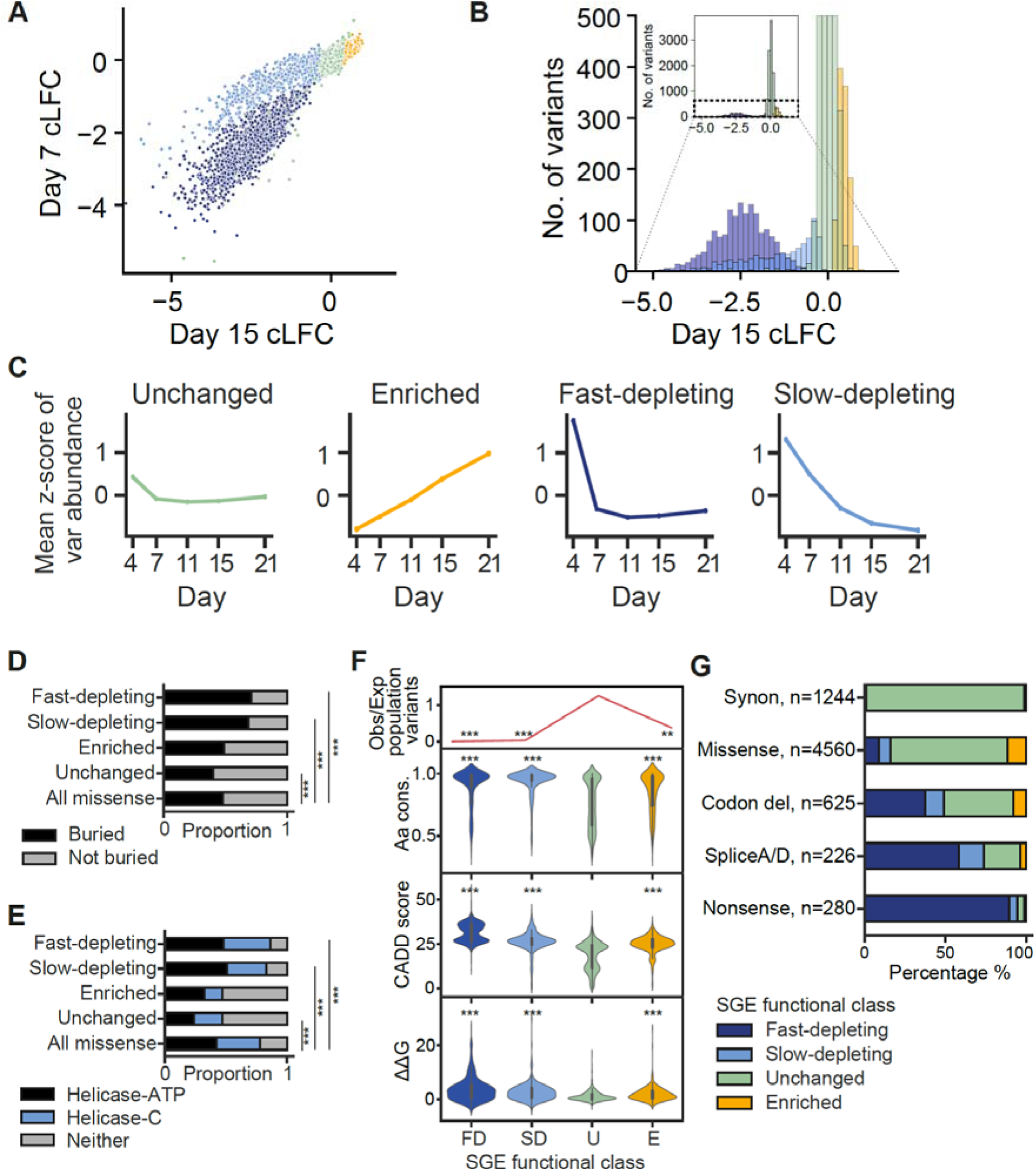
Functional classification of *DDX3X* variants. A/B) *DDX3X* variants with a Day 15 cLFC FDR > 0.01 are considered non-significant/unchanged. Variants with Day 15 cLFC FDR < 0.01 that become more abundant in culture over time are classified as ‘SGE-enriched’. Variants with Day 15 cLFC FDR < 0.01 which become less abundant in culture over time (SGE-depleted) are subjected to a two-dimensional Gaussian Mixture Model using Day 7 and Day 15 cLFC of variant abundance. C) Fast-depleting variants are largely depleted by day 11 in culture while slow-depleting variants deplete more gradually over time in culture. D) Proportion of missense variants that are buried within the protein core (defined as solvent-accessible surface area < 25%) among different SGE functional groups. Compared to all missense variants □^2^ p-value fast-depleting =1.3×10^-19^, slow-depleting = 3.4×10^-13^, SGE-unchanged= 1.4×10^-12^. E) Proportion of missense variants that lie within the helicase domains (UniProt: Helicase ATP-binding domain, hg38: Chr X:41343302-41345442; Helicase C-terminal domain ChrX:41345472-41346968). Compared to all missense variants □^2^ p-value fast-depleting = 2.6×10^-40^, slow-depleting = 5.4×10^-28^, SGE-unchanged = 3.3×10^-12^. F) Top panel: Expected (as estimated by germline neutral mutation rate)/ Observed number of *DDX3X* single nucleotide variants in UKBB and GnomAD databases for each SGE functional class. Fast-depleting (FD) □^2^ p = 7.0 x 10^-12^, slow-depleting (SD) □^2^ p= 2.0 x 10^-8^, SGE-enriched (E) □^2^ p = 0.00024, SGE-unchanged (U) □^2^ p=5.2 x 10^-7^. Second panel: Amino acid conservation scores (Scorecons) for SGE SNV functional variant classes. Kruskal Wallis p=1.1×10^-141^, Dunn’s FDR: fast-depleting = 6.9×10^-81^, slow-depleting = 1.2×10^-66^, SGE-enriched=5.6×10^-24^. Third panel: CADD PHRED scores of SGE SNV functional classes. Kruskal Wallis p=0, Dunn’s post-test BH adj FDR compared to SGE-unchanged: fast-depleting=0, slow-depleting=1×10^-96^, SGE-enriched = 1.42×10^-83^. Lower panel: The change in Gibbs free energy of folding (ΔΔG) for missense variants of different SGE functional classes. Kruskal Wallis p=1.8×10^-54^, Dunn’s post-test BH adj FDR: SGE-enriched=2.4×10^-11^, fast-depleting=3.9×10^-39^, slow-depleting=1.9×10^-23^. Fast and slow-depleting missense variants compared to SGE-enriched variants: Dunn’s post-test BH adj FDR 1.5×10^-6^, 2.3×10^-3^, respectively. G) Proportion of SGE functional classes for synonymous (Synon), missense, codon deletion (Codon del), canonical splice acceptor/donor (SpliceA/D) and nonsense variants across *DDX3X*.

### Properties of depleted and enriched variants

We observed that CADD scores, a widely used *in silico* metric to predict variant deleteriousness^26^, are significantly higher for SGE-enriched and SGE-depleted SNVs compared to SGE-unchanged SNVs (Fig. 2F, bottom panel Kruskal Wallis p=0, Dunn’s post-test BH adj FDR: fast-depleting=0, slow-depleting=1×10^-96^, SGE-enriched = 1.42×10^-83^). Similarly, both SGE-enriched and SGE-depleted SNVs occur at significantly more conserved (higher scorecons score) amino acid positions^27^ than SGE-unchanged SNVs (Fig. 2F, middle panel, Kruskal Wallis p=1.1×10^-141^, Dunn’s FDR: fast-depleting=6.9×10^-81^, slow-depleting=1.2×10^-66^, SGE-enriched=5.6×10^-24^).

A recent study suggested that clinically-relevant loss-of-function variants in haploinsufficient genes are more likely to be observed at residues buried within protein structures, and be more disruptive to protein structure (as predicted by the change in Gibbs free energy of folding (ΔΔG))^28^. We therefore calculated the ΔΔG of missense variants for the different SGE functional classes, and the proportion of these variants that are predicted to be buried within the core of the protein (using relative solvent accessible surface area from structural models)^29^. Missense variants that are fast and slow-depleting are more likely to be observed at buried residues (Fig.2C, □^2^ p=1.3×10^-19^, 3.4×10^-13^, respectively) and within the helicase domains (Fig.2D, □^2^ p=2.6×10^-40^, 5.4×10^-28^ respectively). SGE-enriched, fast and slow-depleting missense variants have a higher ΔΔG than SGE-unchanged missense variants (Fig. 2E, Kruskal Wallis p=1.8×10^-54^, Dunn’s FDR: SGE-enriched = 2.4×10^-11^, fast-depleting=3.9×10^-39^, slow-depleting=1.9×10^-23^). The ΔΔG of fast and slow-depleting missense is significantly higher than that of SGE-enriched variants (Dunn’s FDRs: 1.5×10^-6^, 2.3×10^-3^, respectively).

To investigate whether the different functional classes of variants are likely to be deleterious in human populations, we estimated the number of *DDX3X* variants that we would expect to observe in the Genome Aggregation Database (GnomAD) and UK Biobank (UKBB), based purely on a germline mutation model, assuming no negative selection^30^. SGE-depleted SNVs are observed less frequently than expected in GnomAD and UKBB (Fig.2F, top panel, □^2^ p = 7.0 x 10^-12^, 2.0 x 10^-8^ for fast and slow-depleting SNVs, respectively). This is consistent with loss-of-function variants being highly penetrant for a severe, neurodevelopmental disorder and therefore under-observed in population cohorts. SGE- enriched variants are also observed less frequently than expected in these population variation resources (Fig.2F, top panel, □^2^ p = 0.00024), suggesting that they are also likely to be deleterious, but perhaps less so than SGE-depleted variants.

We attempted to perform a phenome-wide association study in UKBB in order to identify any additional phenotypes associated with the different classes of functionally abnormal *DDX3X* variants that we identified. However, this was hampered by the paucity of individuals carrying SGE-enriched (35 carriers of 16 variants) and SGE-depleted (2 carriers of 2 slow-depleting variants). No phenotypes reached statistical significance after Bonferroni correction for multiple testing (Supplementary Table 7).

### Functional classification of synonymous, nonsynonymous and intronic variants

For simplicity, we focussed downstream analyses on SNVs, excluding the redundant multi-nucleotide variants, which are observed less frequently in humans. We observed that 98% (1271/1301) of synonymous SNVs were SGE-unchanged (Fig.3A, Bi). The 14 SGE-depleted synonymous SNVs include five that are predicted to have a negative impact on splicing by SpliceAI^31^. Three were predicted to be novel splice-donors (SpliceAI delta score for donor gain > 0.9), while two synonymous SNVs were predicted to result in the loss of a splice-donor site (SpliceAI delta score for donor loss = 0.9 and 0.76, respectively).

**Fig. 3.**
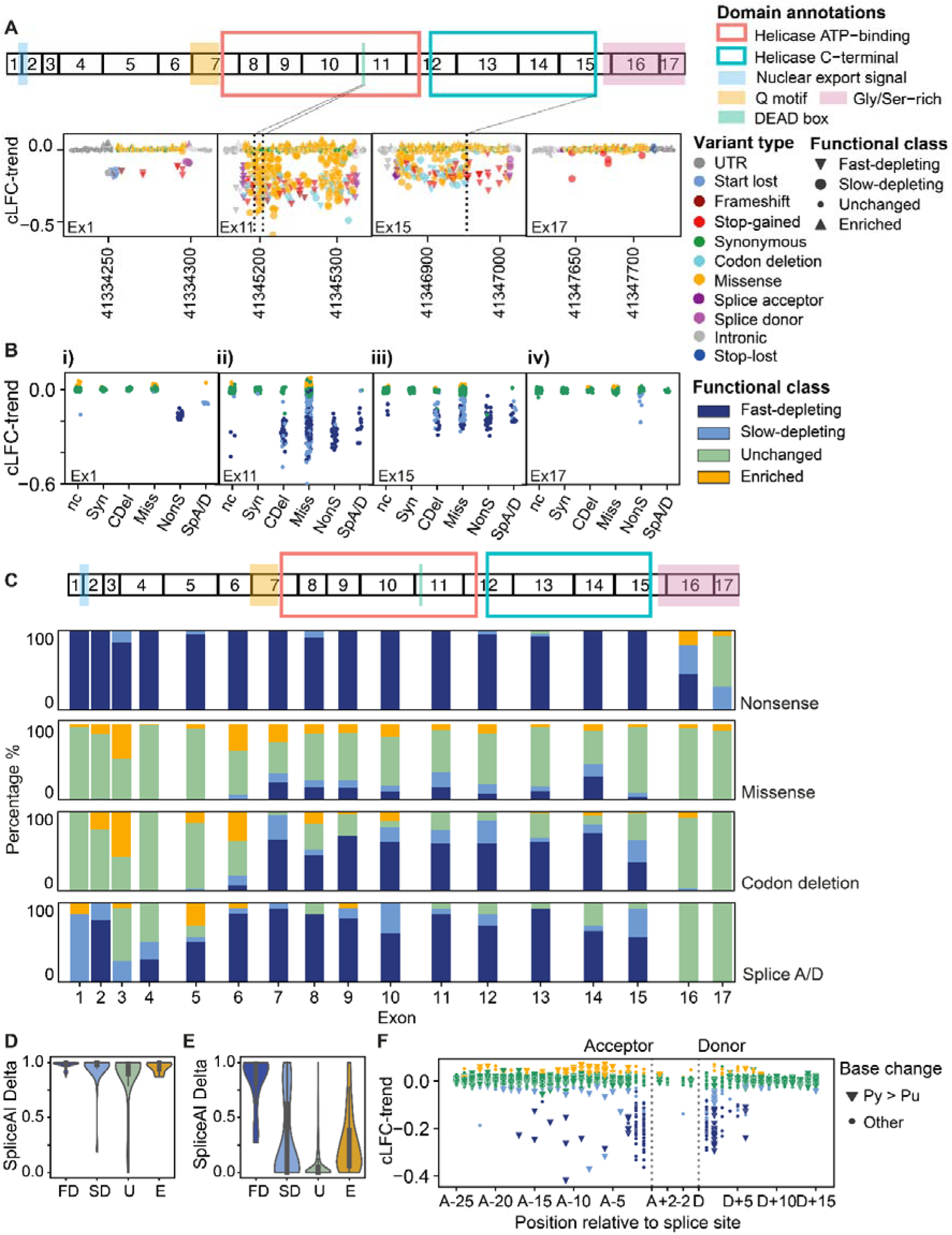
Functional characterisation of *DDX3X* variant types. A) Top panel: Schematic diagram, showing *DDX3X* exons, the position of the helicase ATP-binding, C-terminal domains and Q motif (UniProt annotations), and location of other features such as the DEAD box, C-terminal glycine/serine-rich region and nuclear export signal. Lower panel: y-axis: cLFC-trend; x-axis: hg38 position for variants in exons 1, 11, 15 and 17. B) cLFC-trend of non-coding (nc), synonymous (Syn), in-frame codon-deletion (Cdel), missense (Miss), nonsense (NonS) and canonical splice acceptor/donor variants (SpA/D) in exons 1, 11, 15 and 17, coloured by SGE functional class. C) Proportion of SGE functional classes across exons for nonsense; missense; codon deletion and canonical splice acceptor/donor variants. D) SpliceAI Delta scores for all canonical splice acceptor/donor variants across the gene, by SGE functional class. FD: Fast-depleting, SD: Slow-depleting, U: SGE-unchanged, E: SGE-enriched, E) SpliceAI Delta scores for all intronic variants that are not canonical splice acceptor/donor sites across the gene, by SGE functional class. F) Intronic and synonymous SNVs within 2bp of canonical splice sites grouped according to their position relative to the splice site. Triangles denote pyrimidine to purine variants (Py > Pu), dots all other base changes.

We found that 95% (265/280) of nonsense SNVs were SGE-depleted, including 99.5% (219/220) of nonsense variants predicted to trigger nonsense-mediated decay (NMD)^32^ (Fig.3A, Bi, Bii). Of the 265 SGE-depleted nonsense variants, the majority (95%, 251) were fast-depleting. 7/21 nonsense variants predicted to escape NMD at the 3’ end of the gene were in fact SGE-depleted, 2 fast and 5 slow-depleting (Fig.3A, Bi, Bii). This suggests that some nonsense variants that would be expected to *not* trigger NMD may still have a negative functional effect and are likely of disease-relevance.

Sixteen percent (727/4560) of missense variants are SGE-depleted, 55% of which are fast-depleting (Fig.3A, Bi, Biii). 49% (307/625) of inframe codon-deleting variants are SGE-depleted, 76% of which are fast-depleting. The distribution of SGE-depleted missense and codon-deletion variants is strongly enriched within the previously defined protein domains of DDX3X such as the helicase domains (Fig.2E, 3A, Bii, Extended Data 4; missense and codon-deletion variants: □^2^ p-value 2.1×10^-66^, 7.7×10^-27^, respectively). 92% of all SGE-depleted missense variants occur within the helicase domains and helicase Q motif encoded by exons 7-15. In exon 11, 90% (26/29) of missense variants in the DEAD box domain are SGE-depleted (Fig.3A). In exon 15, we observe an abrupt change in the functional impact of missense and codon-deletion variants which coincides precisely with the UniProt-predicted end of the C-terminal helicase domain (Fig.3A). Throughout the gene, 11.5% (524/4560) of missense variants and 16% (49/625) of codon-deletion variants are SGE-enriched.

74% (167/226) of variants at canonical splice acceptor and donor sites are SGE-depleted, 3.5% (8/226) SGE-enriched and 23% (51/226) unchanged. The proportion of canonical splice sites that are SGE-depleted is notably lower than for nonsense variants. Across the gene, the variants at canonical splice sites that are SGE-unchanged have a significantly lower SpliceAI score than variants that are fast-depleting but not slow-depleting or SGE-enriched (Fig.3D, Kruskal Wallis p=0.002, Dunn’s FDR: fast-depleting=0.001, slow-depleting=0.31, enriched=0.67). Some splice variants can induce exon skipping, although this is difficult to predict computationally^4^. Exon skipping that occurs outside of functionally important domains may produce a smaller protein with preserved function. Exon 3, which is in-frame and lies outside of the functional *DDX3X* domains, has a relative paucity of depleted splice variants compared to out-of-frame exons or exons within the helicase domains (Fig.3C).

Outside the canonical splice donor and acceptor sites, 6% (105/1,886) of intronic variants were SGE-depleted and 6% (104/1886) were SGE-enriched. The SGE-depleted and SGE-enriched intronic variants had higher SpliceAI scores compared to SGE-unchanged intronic variants, however, fast-depleting variants had strikingly higher SpliceAI scores compared to the other SGE functional classes. Intronic sites close to the splice acceptor and donor sites show a bias towards being affected in SGE (Fig.3F). 15% of SNVs (44/288) between donor+3 and donor+8 are affected in SGE, compared to 3% distal to donor +8. This is consistent with previous exome-wide analyses^33^. Within the polypyrimidine tract SGE-enriched and fast-depleting SNVs were significantly enriched for pyrimidine to purine compared to all other base changes (□^2^ p-value fast-depleting =0.006, slow-depleting = 0.35, SGE-enriched= 0.003).

We characterised 87 variants in the 5’ UTR within 25 bp of the start codon. We identified one SGE-depleted UTR variant (ChrX: 41334250 G>T) which would be predicted to generate an upstream start codon, out-of-frame to the native protein.

### Comparison to variants observed in clinical and population cohorts

We identified 536 *DDX3X* coding, canonical splice site and near-exon variants in UKBB or GnomAD population resources, and 239 *DDX3X* variants in individuals with NDD from ClinVar, DECIPHER, the 100,000 Genomes Project and the published literature^1, 20^, not all of which have been clinically interpreted to be pathogenic or likely pathogenic. Variants in NDD individuals represent a mix of variants seen in males and females and include variants with differing inheritance status (*de novo*, inherited, inheritance unknown).

As expected, given the strong negative selection predicted for variants that damage dominant NDD-associated genes^34^, ninety-seven percent (521/536) of variants observed in UKBB and GnomAD were SGE-unchanged, two variants were slow-depleting and 13 were SGE-enriched. By contrast, of the variants seen in NDD cases, 72% (173/239) were SGE-depleted, 3% (8/239) were SGE-enriched and 24% (58/239) were SGE-unchanged (Fig.4A). NDD-observed variants that had been clinically interpreted to be benign were all SGE-unchanged, whereas only 13% (24/181) of those clinically interpreted to be pathogenic or likely-pathogenic were SGE-unchanged, with almost all the remainder (154/181) being SGE-depleted. Using a subset of NDD variants where information on patient sex was available, we noted a strong sex-difference: SGE-unchanged variants are rare among likely pathogenic variants seen in females (12%, 12/117), compared to likely pathogenic variants seen in males (88%, 8/9), consistent with previous studies that suggest that male probands may not carry complete loss-of-function variants^11^ (Fig.4A).

**Fig. 4.**
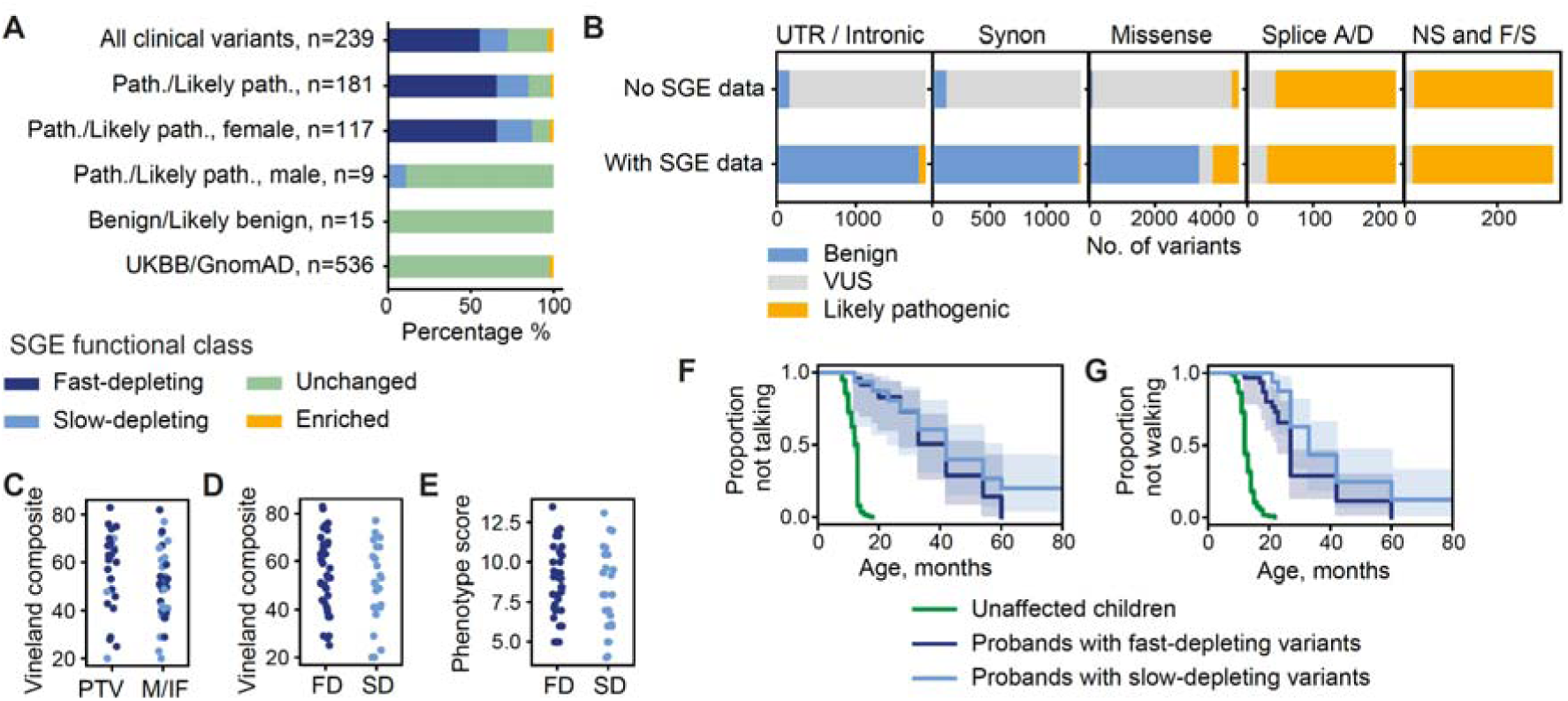
SGE functional classification of *DDX3X* variants observed in clinical and population databases. A) We identified 239 *DDX3X* variants in NDD patients from ClinVar, DECIPHER, the 100,000 Genomes Project and the published literature^1, 20^. SGE functional classification of DDX3X variants, split by clinical interpretation and proband sex (where available); and variants observed in UK Biobank and GnomAD. B) Modelling the impact of SGE data on *DDX3X* clinical variant interpretation in the context of a hypothetical female patient with moderate intellectual disability. Variant classification was performed *in silico* for all possible synonymous, missense, nonsense and canonical splice acceptor/donor (Splice A/D) SNVs and for frameshift (F/S), intronic and UTR variants included in the SGE libraries where the inheritance status of the variant is unknown. The proportion of likely benign/benign, likely pathogenic/pathogenic and VUS are plotted when variant classification is performed with and without incorporating the SGE data. C) Vineland composite scores for individuals with DDX3X-related neurodevelopmental disorder carrying a protein-truncating variant (PTV) or a missense or in-frame variant (M/IF) in three existing clinical phenotyping studies^20,35,36^. D) Vineland composite scores for individuals with DDX3X-related neurodevelopmental disorder carrying fast-depleting and slow-depleting variants. E) Composite phenotypic score for individuals with *DDX3X*-related neurodevelopmental disorder reported by Lennox *et al.* carrying fast-depleting and slow-depleting variants. F) Age at which first words and first independent steps were taken for patients in the DDD study carrying fast-depleting variants and slow-depleting variants, compared to children without a neurodevelopmental disorder. Number of individuals: First words: n=24 fast-depleting variants, n=16 slow-depleting variants; first steps: n=31 fast-depleting variants, n=17 slow-depleting variants.

It is possible to estimate the proportion of all missense variants that are likely to be pathogenic for a given haploinsufficient disease by comparing the relative excesses in large disease cohorts of missense and nonsense *de novo* mutations (compared to the numbers expected under a null germline mutational model). Using the data from over 31,000 NDD families^1^, we estimated that ∼17.5% (95% CI 10.9 - 29.6%) of missense variants in *DDX3X* are likely to be pathogenic for NDDs. This corresponds closely to the 15.9% of missense variants that were found to be SGE-depleted in our assay.

### Training and assessing a machine learning classifier for NDD-relevance

The SGE-based functional classification of *DDX3X* variants described above identified several different functional categories of variants, without utilising clinical information to inform this classification. We reasoned that supervised machine learning of SGE data, trained using clinically interpreted likely pathogenic/pathogenic variants observed in individuals with NDD and putatively benign variants observed in individuals without NDDs, should increase accuracy to identify functionally abnormal variants of NDD-relevance.

We trained a Random Forest machine learning model^37^ using cLFC on Day 7, 11 and 15. Putative truth sets of pathogenic and benign variants were assembled with 80% used for training and the remainder used for testing. The class with the highest mean probability estimate across the trees in the forest model was taken as the model-predicted class.

Model performance was estimated from the ‘test variants’ as 97.1% sensitivity and 100% specificity (Table 1), based on discordant classification of a single variant. As an indicator of variant pathogenicity in a neurodevelopmental context, our NDD classifier has an estimated negative and positive predictive value of 99.1% (Table 1). Clinically interpreted variants may contain some false positives, and, therefore this could be minimum estimates of sensitivity: SGE data outperformed *in silico* predictors of variant effects, which tend to lack specificity at commonly employed thresholds (Supplementary Table 10). Clinically recommended thresholds^38^ prioritise *in silico* predictors’ specificity at the expense of sensitivity (Table 1).

**Table 1.** Evaluation of SGE clinical classifier and comparison to commonly used in silico variant effect prediction algorithms. Minimum thresholds recommended by the ClinGen SVI for use as “supporting” evidence of variant effect within the ACMG variant classification guidelines^38^ for in silico predictors of variant effect were used. ROC AUC = Area under the Receiver-operator characteristic (ROC) curve; TP = the number of test-positive true-positive variants. TN = the number of test-negative true-negative variants. FP = the number of test-positive true-negative variants. FN = the number of test-negative true-positive variants. Sensitivity = 100*(TP/TP+FN), Specificity = 100*(TN/TN+FP), Positive predictive value (PPV) = 100*(TP/TP+FP), Negative predictive value (NPV) = 100*(TN/TN+FN). SGE (RF test vars only) = calculated from truth set variants not employed in the training of the Random Forest classifier for NDD-relevance; SGE (all truth vars) = calculated from all truth set variants, including variants employed for training and testing the Random Forest classifier for NDD-relevance.

|  | ROC<br>AUC | TP | TN | FP | FN | Sensitivity<br>(%) | Specificity<br>(%) | PPV<br>(%) | NPV<br>(%) |
| --- | --- | --- | --- | --- | --- | --- | --- | --- | --- |
| All variants |  |  |  |  |  |  |  |  |  |
| SGE (RF test<br>vars only) | 0.986 | 33 | 108 | 0 | 1 | 97.1 | 100 | 100 | 99.1 |
| SGE (all truth<br>vars) | 0.997 | 167 | 539 | 0 | 1 | 99.4 | 100 | 100 | 99.8 |
| CADD | 0.971 | 108 | 514 | 22 | 21 | 83.7 | 95.9 | 83.1 | 96.1 |
| Missense variants only |  |  |  |  |  |  |  |  |  |
| SGE (RF test<br>vars only) | 0.970 | 16 | 29 | 0 | 1 | 94.1 | 100 | 100 | 96.7 |
| SGE (all truth<br>vars) | 0.995 | 81 | 159 | 0 | 1 | 98.7 | 100 | 100 | 99.4 |
| CADD | 0.898 | 65 | 138 | 21 | 17 | 79.3 | 86.8 | 75.6 | 89.0 |
| REVEL | 0.959 | 21 | 159 | 0 | 61 | 25.6 | 100 | 100 | 72.3 |
| SIFT | 0.927 | 62 | 149 | 10 | 20 | 75.6 | 93.7 | 86.1 | 88.2 |
| Poly Phen2 | 0.916 | 55 | 150 | 9 | 27 | 67.1 | 94.3 | 85.9 | 84.7 |

We then modelled the value that this SGE-based classifier of NDD-relevant functional abnormality might have within existing frameworks for diagnostic variant interpretation. Following the Clinical Genome Resource (ClinGen) Sequence Variant Interpretation (SVI) Working Group recommendations, our SGE-based classifier of NDD-relevant functional abnormality can be used as strong evidence of pathogenic and benign variant effect (PS3 and BS3, respectively^6^, see Methods). *DDX3X-*associated neurodevelopmental disorder has a relatively non-specific phenotype, and usually occurs *de novo* in an X-linked dominant fashion. We therefore estimated what the impact would be of including *DDX3X* SGE data in variant classification in a hypothetical female patient with moderate intellectual disability. Variant classification was performed *in silico* for all possible synonymous, missense, nonsense and canonical splice acceptor/donor variants and for frameshift, intronic and UTR variants included in the SGE libraries in two contexts: where a variant is known to have arisen *de novo* ( Supplementary Table 10) and where the inheritance status of the variant is unknown^3, 4, 6, 39^ (Fig. 4C, Supplementary Table 11). The number of missense VUS of unknown inheritance was reduced by 90%, from 4,261 to 423 variants, with a nearly four-fold increase in the number of likely pathogenic/pathogenic variants (199 to 779) and an over 30-fold increase in the number of likely benign/benign variants (100 to 3,358; Supplementary Table 11). For synonymous and non-coding variants, incorporation of SGE data reduced the number of VUS by over 99%, with the majority of variants being reclassified as benign. However, 15 synonymous and 74 non-coding (intronic and UTR) variants were reclassified as likely pathogenic/pathogenic (Fig.4C, Supplementary Table 11).

Unsurprisingly, our SGE assay had much less effect on the classification of predicted loss-of-function variants such as nonsense, frameshift and canonical splice-site variants. This is partly because of the high concordance between predicted loss-of-function annotation and empirical evidence of functional abnormality in our SGE assay, but also because of the substantial weighting accorded to these variant annotations in the ACMG guidelines^3, 4^, such that functional data that supports a benign interpretation can be outweighed. For example, under the existing guidelines, a canonical splice variant predicted to trigger NMD which has not been previously reported in either patient or healthy populations would remain likely pathogenic even after the incorporation of SGE data which suggests that the variant remains functional, thus supporting a benign interpretation.

To exemplify the real-world use of this NDD-relevant functional classifier, we applied it to 23 rare nonsynonymous variants in *DDX3X* of unknown inheritance status (due to the lack of DNA from both parents) from the DDD study^40^. We found that 39% (9/23) of these variants were classified as being functionally abnormal, and 61% as functionally normal. 8/9 of functionally abnormal variants were in female probands.

### Genotype-phenotype correlation

To investigate whether there are differences in the severity of intellectual disability between DDX3X-related NDD probands who carry SGE-fast and SGE-slow-depleting variants we identified 61 probands who had undergone assessment via the Vineland Adaptive Behaviour Scales^41, 42^ across three studies^20, 35, 36^. No difference in global adaptive function (Vineland Adaptive Behavior Composite score) was observed between individuals carrying missense variants and those carrying protein-truncating variants (PTVs) (Fig.4C). There was also no significant difference in the global adaptive function of individuals carrying fast or slow-depleting *DDX3X* variants (t-test p=0.27, Fig. 4D). To investigate phenotypes more broadly, a composite score was devised for the Lennox *et al.* cohort, encompassing brain MRI findings, microcephaly, sensory deficits, muscle tone anomalies, cardiac findings, precocious puberty, experience of seizures and behavioural assessment. We observed no significant difference in composite score between fast and slow-depleting variants (Fig.4E). Finally, we observed no difference in the rate of attainment of developmental milestones (age of speaking first words or taking first independent steps) in patients in the DDD cohort carrying fast or slow-depleting variants (Fig.4F).

### *DDX3X* in Cancer

Putative *DDX3X* driver variants have been reported in diverse cancer types including medulloblastoma^15–17^, various lymphoid tumours^43–48^, and melanoma^49^. However, the role of *DDX3X* in oncogenesis remains unclear, as it has been classified as both a tumour suppressor and an oncogene^15, 18, 19, 50^. In cancer types where *DDX3X* does not act as a driver gene, one would expect the proportions of *DDX3X* variants in the different SGE functional classes to be driven solely by the underlying somatic mutational processes. If *DDX3X* acts as a tumour suppressor gene, we would expect to observe an increased proportion of SGE-depleting variants, as these loss-of-function variants would confer a growth advantage. To assess this, we analysed a dataset from cBioPortal^51, 52^ comprising *DDX3X* nonsynonymous variants identified in 90,279 cancer samples. Variants were stratified according to whether *DDX3X* has been identified as a putative driver gene^53^ in the cancer-type of origin. The proportion of fast and slow-depleting variants in *DDX3X*-driver cancers is 2.5 and 2.4x greater than in *DDX3X-*non-driver cancers (□^2^ p= 9.0 ×10^-5^, 9.6 ×10^-5^, respectively), while the proportion of SGE-enriched variants observed in these cancers is not significantly different (□^2^ p = 0.41) (Fig. 5A).

**Fig. 5.**
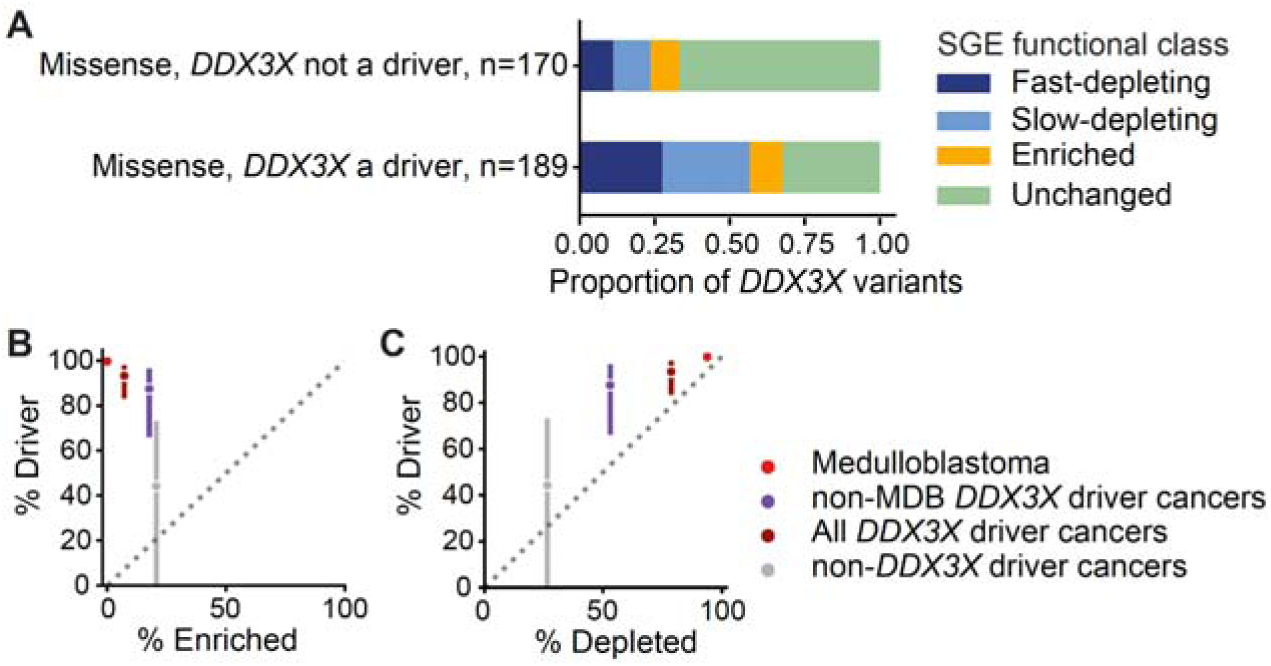
SGE functional classification of *DDX3X* variants observed in cancers. A) *DDX3X* missense variants observed in 90,279 cancer samples from cBioPortal were split depending on whether or not *DDX3X* had been identified as a putative driver gene in each cancer type. The proportion of fast-depleting and slow-depleting variants in cancer types where *DDX3X* has been identified as a driver gene is 2.5 and 2.4x greater than in cancer types where *DDX3X* has not been identified as a driver (□^2^ p=9.0 ×10^-5^, 9.6 ×10^-5^, respectively); while the proportion of SGE-enriched variants observed in these cancer types is not significantly different (□^2^ p=0.41). B/C) y-axis: The percentage of *DDX3X* missense variants predicted to be driver variants by dNdScv; x-axis: the percentage of *DDX3X* missense variants that are B) SGE-enriched, C) SGE-depleted. Scarlet: medulloblastoma tumours only, Purple: cancer types excluding medulloblastoma (MDB) in which *DDX3X* was identified as a driver gene by dNdScv, Dark red: all cancer types in which *DDX3X* was identified as a driver gene by dNdScv, Grey: cancer types in which *DDX3X* was not identified as a driver gene by dNdScv.

To further investigate the role of *DDX3X* variation in cancer, we used dNdScv, for the subset of cancers for which synonymous variant data are available^54^, to estimate the proportion of *DDX3X* missense variants acting as drivers in different cancer types, and to test how well this correlates with the proportion of these variants that appear functionally abnormal in our SGE assay, while correcting for mutational differences between cancers. We analysed mutational profiles of 33 different cancer types from The Cancer Genome Atlas (TCGA) and the Pan-Cancer Analysis of Whole Genomes^55^. We found that the proportion of missense somatic mutations estimated to be driver variants was highly correlated with the proportion of those variants that are SGE-depleted, but not with the proportion that was SGE-enriched (Fig. 5B,C). We observed that missense somatic mutations that were SGE-depleted could almost entirely account for the number of expected driver mutations per cancer. Notably, in medulloblastoma 99.7% (95% CI 99.1-100) of *DDX3X* missense variants are estimated to be drivers, and all are SGE-depleted (n=16). Together, these data support the argument that *DDX3X* predominantly acts as a tumour suppressor gene across multiple tumour types.

## Discussion

We have described a near-exhaustive characterisation of exonic and near-exonic variant effects for the gene *DDX3X*, comprising 12,776 variants across 17 coding exons. Studying multiple time points of cell culture enabled us to identify three classes of functionally abnormal variants: rapidly-depleting, slowly-depleting and enriched. This map corresponds very well to the predicted protein consequence of variants, with almost all nonsense variants being functionally-abnormal and almost all synonymous variants being functionally-normal. Nonetheless, this map includes tens of functionally-abnormal variants that might otherwise be expected to be functionally-normal (e.g. synonymous, intronic and UTR variants) and tens of functionally-normal variants that might have been expected to be functionally-abnormal (e.g. over 20% of variants at canonical splice sites).

We trained a machine learning classifier to identify functionally-abnormal variants of NDD-relevance. This had much higher accuracy in discriminating between pathogenic and benign variants than a range of *in silico* tools in common usage for clinical variant interpretation, and provides strong enough evidence to resolve the majority of potential VUS in *DDX3X* using the existing ACMG guidelines (Supplementary Table 10 and 11). Our classifier of functionally-abnormal variants of NDD-relevance only classifies one (out of 168) *de novo* pathogenic/likely pathogenic variant observed in a female with NDD as being functionally-normal (i.e. not depleted in the SGE) at hg38 ChrX:41343346, A>G, NP_001347.3:p.Q225R. This missense variant was deposited to ClinVar (ClinVar ID 975674) in 2020 as a pathogenic *de novo* variant, interpreted according to the 2015 ACMG guidelines^3^, but no criteria have been provided to support this assertion. This variant would likely be interpreted as a VUS following the 2018 ClinGen revised variant interpretation guidelines^4^.

Functionally-abnormal variants that were depleted in our assay were significantly over-represented in cancer types in which *DDX3X* has been shown to be a driver gene, whereas functionally-abnormal variants that were enriched in our assay were not. Indeed this over-representation of functionally-abnormal depleted variants could account for almost all of the excess nonsynonymous *DDX3X* somatic mutations seen in these cancers. This suggests, firstly, that the vast majority of cancer driver mutations in *DDX3X* are loss-of-function variants, and that *DDX3X* predominantly operates as a tumour suppressor gene, and, secondly, that this functional classification of variant effect identifies almost all of the cancer driver mutations. These findings should help to resolve the ongoing uncertainty around *DDX3X*’s mechanism of action in cancer^15, 18^. Moreover, we noted that functionally-abnormal depleted variants in *DDX3X* are also over-represented, albeit more modestly, in cancer types in which *DDX3X* has not yet been conclusively demonstrated to be a cancer driver gene. This suggests that *DDX3X* plays a tumour suppressor role in a wider range of cancers than currently appreciated, and that integration of this variant effect map into analyses of somatic mutation enrichment should increase power to identify other cancers in which *DDX3X* plays a role.

We noted a striking difference between the concordance of the variant effect map with variants clinically interpreted to be pathogenic or likely pathogenic in females with NDD, compared to the almost complete discordance with variants clinically interpreted to be pathogenic or likely pathogenic in males with NDD. We consider three possible explanations for this: (i) the ‘pathogenic’ variants in males have been mis-interpreted and are not actually disease-relevant, (ii) the ‘pathogenic’ variants in males are pathogenic and operate via a mechanism probed in our assay, but our assay is insufficiently sensitive to detect their more subtle functional effects, and (iii) the ‘pathogenic’ variants in males are pathogenic but operate via a mechanism that is not probed by our assay (e.g. could be cell-type specific). We note that Martin *et al.* previously reported that the statistical evidence for males with NDDs having an excess of nonsynonymous rare variants in *DDX3X* compared to males without NDDs is modest, and would not survive Bonferroni correction for multiple testing of all genes on the X chromosome, let alone genome-wide. This was despite the large sample sizes and statistical power of their study. Martin *et al.* found 12 rare missense *DDX3X* variants in 7,844 males with NDDs compared to 11 rare missense *DDX3X* variants in 8,551 males without NDD, suggesting, at the very least, that the majority of rare missense variants in *DDX3X* in males with NDD will not be pathogenic^14^. The concordance between these prior statistical genetic analyses and the functional data presented here with regard to the lack of strong evidence for a pathogenic role of *DDX3X* variants in males with NDD suggests that further evidence is needed before concluding that damaging variants in *DDX3X* can cause NDDs in males. This evidence could comprise rigorous statistical genetic assessment of the evidence that damaging missense variants in *DDX3X* are significantly over-represented in males with NDDs, as well as functional evidence that putatively pathogenic missense variants in males share mechanistic, and potentially phenotypic, similarities that are distinct from pathogenic variants seen in both females with NDDs and in cancer.

Relatively few *DDX3X* variants have been functionally characterised previously, against which we could compare our results. Fonseca *et. al.* proposed that the L556S and R376C *de novo* variants observed in female patients render DDX3X prone to protein aggregation^22^. Both variants are functionally abnormal depleting variants in our assay. Kellaris *et al.* modelled *in vivo* in zebrafish embryos the R79K variant seen in two male siblings with NDD and proposed that this variant results in a partial loss-of-function^21^. This variant is functionally normal in our assay. Snijders-Blok *et al.*^11^ applied the same *in vivo* assay to assess 3 variants observed in male probands and 5 observed in females and observed all male variants to be no different to wild-type. Of the 5 female variants for which they identified loss-of-function effects (I214T, R326H, R376C, I507T, R534H), 4 are functionally abnormal depleting variants in our assay. Of the 8 variants previously determined to have negatively impacted helicase activity^20^, 7 are functionally abnormal depleting variants in our assay. The remaining variant, R326H, was interpreted to be pathogenic and functionally abnormal by both Lennox and Snijders Blok *et al.*. The SNV generating R326H in our assay is classified as functionally normal. However, the multi-nucleotide redundant variant for the same amino-acid change is a functionally abnormal, fast-depleting variant.

We identified functionally-abnormal *DDX3X* variants with different kinetics of depletion *in vitro*. Cells carrying slow-depleting variants persist for longer in culture than those carrying fast-depleting variants. 95% of nonsense variants are fast-depleting compared to 55% of missense variants. Therefore, it is possible that the rate of depletion in culture reflects differing extents of loss of *DDX3X* function. However, we were not able to discern any phenotypic differences between patients carrying fast and slow-depleting variants, suggesting that different kinetics in short-term culture may not be biologically meaningful in the context of organismal neurodevelopment over years. Alternatively, we may lack power to discern such effects, as most phenotyping studies remain small, employ disparate neuropsychometric tests and not all patients are uniformly assessed^11, 20, 35, 36^. It is also notable that *DDX3X* escapes X-inactivation to a variable degree between individuals^56–58^, and a high incidence of severe skewing of X-inactivation has been observed in patients with *DDX3X*-related neurodevelopmental disorder^11, 22^. Such factors will affect variant expressivity and may obfuscate identification of genotype-phenotype relationships. Previous studies have suggested that patients with missense variants in *DDX3X* have reduced adaptive ability and a more severe clinical phenotype than those with protein-truncating variants^20, 36^. However, in this larger meta-analysis, we see no evidence for different adaptive ability between patients carrying missense versus PTV variants (Fig.4Di).

This study represents the largest variant effect map yet produced by SGE, larger than those of *BRCA1* (3,893 SNVs) and *CARD11* (2,542 variants)^7, 59^, and the only one to include all coding exons. We modified the previously published SGE protocol in HAP1 cells, utilising two independent sgRNAs and HDR libraries per exon, improving transfection efficiency, using a Cas9 expressing HAP1 cell line, including a broader range of variants within HDR libraries and including a time-course of sampling over a longer culture. These changes enhance the sensitivity of the SGE assay. Inclusion of a time-course allowed us to identify both loss-of-function variants which have different kinetics of variant effect (fast- and slow- depleting) and a subset of *DDX3X* variants (enriched) which do not appear to act through a loss-of-function mechanism, but which are rarely observed in healthy individuals. We included a more systematic screen of indels than in previous SGE studies, including codon deletions for all contiguous codons. The pattern of functionally abnormal codon deletions mirrors the predicted domain structure of the protein and should assist in the clinical interpretation of in-frame indels.

All three SGE studies^7, 59^ have shown that a MAVE is more specific than *in silico* metrics in discriminating between pathogenic and benign variants. This demonstrates the limitations of evolutionary constraint-based metrics which cannot distinguish between different mechanisms of variant effect to identify disease-relevant variant subtypes. This may explain why *in silico* metrics of variant effect often have high sensitivity but limited specificity, (Table 1, Supplementary Table 10)^38^. MAVEs are therefore likely to be of particular value when applied to genes where purifying selection results from different mechanisms, not all of which are relevant for a specific disease.

HAP1 cells are derived from the chronic myelogenous leukaemia line KBM-7 by introducing the Yamanaka factors^60^, and, as a cell type, lack apparent pathophysiological relevance to NDDs. Here, HAP1 cells in SGE are being used as a platform for the high-throughput assessment of variant impact on gene function, and not as a model of pathophysiology. The fact that a functional assay in such an unnatural cell type is of demonstrable value for the interpretation of variant effects in NDDs and cancer suggests that, for some genes at least, the functional impact of variation is predominantly protein-intrinsic and cell-type-agnostic rather than cell-context specific. It remains to be seen for what fraction of disease-associated genes will pathogenic variation be predominantly protein-intrinsic. With readily accessible databases of pathogenic and benign variation, it is possible to validate the utility of variant effect maps empirically and not rely on biological intuition or expert consensus^6^ with regard to the *a prior*i relevance of a given model system.

Further investigation is needed into the functionally abnormal enriched variants, to establish the mechanism of action of these variants, what their phenotypic consequences might be, and how this results in purifying selection acting on these variants.

Finally, having demonstrated the utility of a single variant effect map in an experimentally tractable cell type across distinct conditions (here NDD and cancer), we are excited and motivated by the opportunity to scale up the generation of useful variant effect maps for thousands of disease-associated genes. There are 278 genes associated with developmental disorders or cancer (DDG2P - www.ebi.ac.uk/gene2phenotype ; Cancer Gene Census - https://cancer.sanger.ac.uk/census ) that have a pathophysiological loss-of-function mechanism and appear to be essential for HAP1 cell growth^23, 24^. Experimental refinements will be needed to increase the scalability of SGE, however, this will be a collective, international endeavour (www.varianteffect.org) and open sharing of resources, protocols, code and data will propel progress.

## Data Availability

The sequencing data generated in this study were deposited to ENA with accession number PRJEB52929.

https://www.ebi.ac.uk/ena/browser/view/PRJEB52929

https://www.cancer.gov/tcga

https://www.ncbi.nlm.nih.gov/clinvar/docs/maintenance_use/#download

https://gnomad.broadinstitute.org/downloads

https://www.deciphergenomics.org/

## Acknowledgements

This work was supported by core Wellcome funding to the Wellcome Sanger Institute (grant Grant reference number: 108413/A/15/D), and for the purpose of open access, the author has applied a CC BY public copyright licence to any Author Accepted Manuscript version arising from this submission. This work was also funded by a Clinical Lecturer Starter Grant from the Academy of Medical Sciences, the Wellcome Trust, the Medical Research Council, the British Heart Foundation, Versus Arthritis, Diabetes UK, the British Thoracic Society (Helen and Andrew Douglas bequest), and the Association of Physicians of Great Britain and Ireland to E.J.R. [SGL023\1060]. E.J.G. and J.R.B.P. are funded by the Medical Research Council (Unit programs: MC_UU_12015/2, MC_UU_00006/2, MC_UU_12015/1, and MC_UU_00006/1). We thank the Cytometry Core Facility, Molecular Cytogenetic Team and High Throughput Sequencing Team at Wellcome Sanger Institute for the technical support in this study. We thank E. Delage for assistance with data deposition. We also thank L. Parts, N. Whiffin, G. Findlay, M. Gasperini, K. Samocha, J. Kaplanis, A. Bassett, J. Buxbaum, D. Grice, E. Sherr and members of the Hurles group and AVE alliance for useful discussion on data analysis. We thank F. Day for assistance in preparing cancer phenotype data derived from the UK Biobank. This research was made possible through access to the data and findings generated by the 100,000 Genomes Project. The 100,000 Genomes Project is managed by Genomics England Limited (a wholly owned company of the Department of Health and Social Care). The 100,000 Genomes Project is funded by the National Institute for Health Research and NHS England. Wellcome, Cancer Research UK and the Medical Research Council have also funded research infrastructure. The 100,000 Genomes Project uses data provided by patients and collected by the National Health Service as part of their care and support. The DDD study presents independent research commissioned by the Health Innovation Challenge Fund (grant no. HICF-1009-003). The full acknowledgements can be found online (www.ddduk.org/access.html). The results used in the analysis are in part based upon data generated by the TCGA Research Network: https://www.cancer.gov/tcga. PheWAS analyses were conducted using UKBiobank application 9905 (to J.R.B.P.). We thank the DDX3X Foundation and DDX3X Support UK for their engagement with and enthusiasm for this research. This study makes use of data generated by the DECIPHER community. A full list of centres who contributed to the generation of the data is available from https://deciphergenomics.org/about/stats and via email from. Funding for the DECIPHER project was provided by Wellcome.

## Data availability

The sequencing data generated in this study were deposited to ENA with accession number PRJEB52929.

## Code availability

All codes used in this study are available from the Github repository (https://github.com/HurlesGroupSanger/Saturation_Genome_Editing/tree/main/Codes).

## Methods

### SGE HDR oligo design and variant annotation

The HDR-library oligos (Supplementary Files 1 and 2) were designed with a custom script. In short, the coding sequence of each exon and its 25bp upstream and 15bp downstream were extracted from the hg38 reference genome. Transcript NM_001356.4 was used for the design of the codon-related variants. Synonymous mutations at the sgRNA protospacer and PAM were then introduced manually to the extracted sequences. Then, the following oligonucleotides were designed: every SNV at every base position; tiled two base-pair deletions in the UTRs and intronic regions; in-frame deletion of every possible codon; and at least one redundant codon for each SNV at each base position, if possible. By redundant codon, we mean an alternative triplet code for each designed SNV. The redundant codons are therefore multinucleotide variants. All indels shorter than 50bp observed in these regions in GnomAD, ClinVar or DECIPHER were added manually to the final library pool. Illumina P5/P7 sequence and the homology sequences for Gibson cloning were appended at both ends of the oligos. All designed HDR-library oligos had at most 300 bases and were synthesized by TWIST Bioscience. HDR-library-associated variant call format (VCF) files were generated by re-running the same design with VaLiAnT^61^. VaLiAnT VCFs were then used to generate annotation files through VEP .

### TWIST oligo library preparation

The step-by-step protocols used in this study were deposited to Github (https://github.com/HurlesGroupSanger/Saturation_Genome_Editing/tree/main/Wetlab_protocols). The TWIST HDR-library oligo pools, consisting of designed HDR-library oligos from 17 exons, were dissolved in water at 10ng/uL. 100ng of the HDR-library oligo pools were amplified by P5 and P7 primers for 18 cycles with an annealing temperature of 60°C in 2 X 100 uL reactions. The designed variant HDR libraries of each exon were then extracted from the amplified pools by PCR with exon-specific primers. The primer sequences for each exon of *DDX3X* are listed in Supplementary Table1. All primer oligos used in this study were purchased from IDT. 2X KAPA HiFi Hotstart ReadyMix (KAPA Biosystems) was used for all PCR reactions in this study, according to the manufacturer’s protocol.

### Plasmid preparation

pMin-U6-ccdb-hPGK-puro (Supplementary File 3) was used for both sgRNA plasmid and HDR template library plasmid constructs. For sgRNA cloning, the annealed sgRNA oligo was cloned into the BbsI-HF (NEB) digested pMin plasmid. For HDR template library plasmid cloning, a genomic sequence of 800bp upstream and downstream of a *DDX3X* exon was first amplified from HAP1 gDNA and followed by cloning with Gibson assembly (NEB) into the NotI-HF (NEB) and SbfI-HF (NEB) digested pMin plasmid. The homology-arm plasmid sequences were validated by Sanger sequencing (Eurofins). The validated plasmid was linearized by PCR and was assembled with the TWIST HDR template library by NEBuilder® HiFi DNA Assembly kit (NEB). The reaction was then transformed into Stellar chemically competent cells (Takara) according to the manufacturer’s protocol. The HDR plasmid libraries were subjected to maxiprep (Qiagen) and quality control by Illumina Miseq with 2 X 280 pair-end sequencing. The sgRNA oligo sequences are listed in Supplementary Table 2. Primary sequences for preparation of the HDR template libraries are listed in Supplementary Table 3.

### HAP1 cell bank preparation

The HAP1 *LIG4* knock-out (KO) clone, HZGHC000759c005, which carries a 10-base deletion at the *LIG4* exon 3 gene locus (hg38: Chr13:108,210,833-108,210,842) (Extended Data 1C), was purchased from Horizon Discovery and cultured according to the manufacturer’s protocol. This HAP1 line was then transduced with the pKLV2-EF1a-BsdCas9-W (Addgene, 67978) lentivirus and was selected in 10ug/mL blasticidin (Gibco) to generate *LIG4*-KO Cas9-expressing HAP1 cells. The Cas9 positive cells were then stained with 10ug/mL of Hoestch 33342 (ThermoFisher Scientific) followed by sorting of the haploid G1 population with an XDP FACS sorter. The sorted haploid cells were expanded in the presence of 10ug/mL of blasticidin for one week and were then banked in liquid nitrogen. The cells were sent for mycoplasma tests and karyotyping. (Extended Data 1D)

### Ploidy assay

HAP1 cells were arrested at metaphase by treatment with 0.1uM nocodazole (Biovision) for 14 hours in a 37°C incubator. The treated cells were then dissociated with TrypLE (Thermo Fisher) and fixed with 80% ethanol. The fixed cells were treated with 0.1% TritonX-100 in PBS. 5×10^5^ cells were then resuspended in 500uL of 0.1% TritonX-100/PBS with DAPI at 1ug/mL and incubated at room temperature for 30 minutes before performing the FACS assay. Singlet cells were gated for the analysis. Haploid and diploid cell populations were separated using the DAPI channel. (Extended Data 1E)

### Cell culture and cell transfection

HAP1 cells were thawed and expanded one week before each screen started. 8×10^5^ of HAP1 cells were seeded in each well of a 12-well plate one day before transfection (Day -1). 6 to 12 wells were used for each biological replicate in the SGE. 2ug of the sgRNA plasmid and 4ug of the HDR template library plasmid were transfected into each well with 3.6uL of Xfect transfection reagent and 100uL of Xfect buffer (Takara). Media change was performed after 4 hours of incubation at 37C. The cells were selected with 3ug/mL of puromycin (Gibco) for two days, from 24 to 72 hours after transfection (Day 1 and Day 2). The cells were passaged on Day 3 and were seeded into two T75 flasks. One of the flasks was harvested on Day 4, and the other flask was passaged on Day 5. The cells were then passaged every two days by re-seeding 10-25 million cells in 1-3 T150 flasks until Day 21. Cell pellets were harvested on Days 7,11,15 and 21. The cells were maintained with at least 10,000X library coverage for every passage.

### Sequencing library preparation

HAP1 genomic DNA (gDNA) was harvested by Qiagen DNeasy Blood and Tissue kit or Qiagen AllPrep DNA/RNA mini kit. 3-step nested PCR was performed to amplify the targeted exon from the HAP1 gDNA. To avoid amplifying the HDR template library plasmid that may persist transiently in the cell culture, the first-step PCR primers were designed outside the homology arm sequences of the targeted exons. For the second-step PCR, Illumina adaptor sequences were appended at the 5’ end of the target-specific primer sequences. Illumina P5/P7 and index sequences were added during the third-step PCR. 4ug of gDNA was used for the first-step PCR (4 x 50uL reaction). 100ng of the first-step PCR product was used for the second-step PCR (4 x 50uL reaction), and 25ng of the second-step PCR product was used for the third-step PCR (1 x 50uL reaction, 7 cycles). The PCR reactions were purified with QIAquick 96 PCR Purification Kit (Qiagen) or with AMPure XP bead (Beckman Coulter) and the PCR products were quantified either by nanodrop or by Qubit dsDNA HS assay kit (Thermo Fisher). The pooled library was quantified with KAPA Library Quantification Kit Illumina® Platforms (KAPA Biosystems) and was sequenced on an Illumina Hiseq 2500 Rapid platform with 1 x 300 single-end. The primers used for each exon’s sequencing library preparation and its PCR cycling conditions are listed in Supplementary Table 4.

### Calculation of variant abundance, Log Fold Change (LFC) and LFC trend

Illumina adaptor sequences in the raw fastq file were removed by Trim-galore.^63^ The exon sequences were then extracted with Tagdust2 ^64^ by using the primer sequences as the adaptor sequences. Each of the unique sequences was counted and the count tables were processed by R. For each sgRNA/HDR template library, the count table of each replicate and timepoint were joined. The joined tables were used for the DESeq2 analysis.^25^ Sequences with a total read count less than or equal to 10 were removed from the analysis. The default DESeq2 scaling factor was replaced by each sample’s total count-normalization factor. Day4 was used as a reference baseline for the LFC calculation of each time point. For the LFC trend calculation, the same DESeq2 code was applied, except that the timepoints were changed to “numeric”, such that the baseline Day 4 was set to “0” and Day 7,11,15,21 were set as “3”,”7”,”11”,”17” respectively. The sequences that matched designed oligos were retained for the subsequent analysis. The median of the LFC/LFC-trend from the sequences annotated as “synonymous_variant” and “intronic_variant” (by VEP) were subtracted from the LFC/LFC-trend of each variant. To combine the results (calculate for cLFC/cLFC-trend) from two independent sgRNA screens within the same exon, Inverse-Variance-Weighted Average and Weighted Sum of Z-Scores were applied to the LFC/LFC-trend where the sequences referred to the same DNA variant.^65^

### Analysis of Gibbs free energy of folding (ΔΔG)

Gibbs free energy of folding (ΔΔG) values for DDX3X was obtained from Mutfunc^66^. ΔΔG was calculated using FoldX from the crystal structure of human DDX3X (PDB 5e7i). DDX3X ΔΔG values are provided in Supplementary Table 5

### Determining whether amino acid residues are buried versus exposed

The total solvent accessible surface area (units: Angstrom^2) (SASA), and the surface area of each residue was obtained from the POPScomp server (http://popscomp.org:3838/) for the following PDB structures: 5e7i, 5e7j, 2jgn, 6o5f, 4px9, 2i4i, 6cz5, 5e7m, 4pxa, and the alpha-fold predicted structure of DDX3X: AFO00571^29^. SASA values were only considered between amino acid residues 133 and 585, as data were available from at least 2 structures across this region. The mean SASA and mean residue surface area across these structures were calculated. Then the proportion of accessible surface area was calculated (mean SASA/ mean residue surface area). A residue was considered to be “buried” if the proportion of accessible surface area was less than 25%. The proportion of accessible surface area values for DDX3X amino acids are given in Supplementary Table 5.

### Amino acid conservation scores

Conservation scores for each amino acid in DDX3X were generated as follows: BLASTP^67^ was run on the protein sequence of human DDX3X (UniProt ID O00571) against the UniRef90 database with the following parameters: *--matrix BLOSUM62 --exp 10 --dropoff 0 --alignments 1000 --scores 1000 --gapopen 10 --gapext 1 --align 0 --filter F --async*. In order to limit the amount of processing, 200 pairwise alignments with a minimum identity of 25% were selected (Supplementary File 4). A pile-up alignment was built, inserting gaps where necessary, using O00571 as the reference sequence. The Scorecons algorithm^27^ (https://www.ebi.ac.uk/thornton-srv/databases/cgi-bin/valdar/scorecons_server.pl), was used to compute the residue conservation scores with the following modification: where a matched sequence is shorter than the search sequence the “gaps” at the N- and/or C-terminal ends were excluded. Scorecons conservation scores, and the amino acid variation at each residue position, are given in Supplementary File 5.

### Clinical variant curation

Clinical variants identified in the context of *DDX3X*-related neurodevelopmental syndrome were identified from the following sources: ClinVar (4th December 2020); DECIPHER (4th December 2020); Genomics England 100,000 genomes study (21st January 2021); and the literature ^1, 20^. Benign and likely benign variants were grouped together. To generate a high-confidence set of likely pathogenic *de novo* variants, the following variants were included; ClinVar: *de novo* variants annotated as likely pathogenic or pathogenic, for which there were no conflicting interpretations; DECIPHER: *de novo* variants annotated as likely pathogenic or pathogenic; Genomics England 100,000 genomes study: *de novo* variants considered clinically relevant (Tier 1 and 2 variants); *de novo* variants identified in a recent meta-analysis of 31,000 developmental disorder exomes considered clinically relevant, and where these variants had not been clinically interpreted as of uncertain significance or benign^1^; *de novo* likely pathogenic or pathogenic variants reported in Lennox *et al.*^20^. As the information on proband’s sex is not available for all of these variants, no filtering based on proband sex was performed. If a variant was curated into the list of *de novo* likely pathogenic/pathogenic variants, it was included, even if it was observed as an inherited variant or a VUS in another source. Variants were removed from the list of VUS if they had been reported as likely pathogenic/pathogenic or likely benign/benign by another source. See Supplementary Table 6 for all clinical variants, together with information on their source and clinical interpretation. Where the same variant is observed in multiple repositories it is not possible to ascertain whether the variant is observed in the same or different probands.

### GnomAD and UKBB variant curation and PheWAS analysis

GnomAD *DDX3X* variants were downloaded on 21st January 2021. Variants from GnomAD v2.1.1 were lifted over to build hg38 using python liftover (https://github.com/jeremymcrae/liftover). UKBB: data from all 454,787 individuals with available whole-exome sequencing from the UKBB was used for PheWAS analysis. For all other analyses, the previous 200,629 individual whole-exome dataset was used.^68^

To identify variants in *DDX3X* observed among the UKBB^69^ individuals, we queried GRCh38-aligned population-level VCF provided via the UKBB research access platform (showcase field 23148). Prior to analysis, we split multiallelic variants and left-corrected and normalised indel variants using bcftools.^70^ We next applied genotype-level filtering where individual genotypes were set to null (i.e. “./.”) if the genotype had a depth < 7, genotype quality < 20, was called as heterozygous in a genetically male individual, or a binomial test p-value for alternate versus reference reads for only heterozygous genotypes ≤ 0.001. If more than 50% of genotypes were missing for a given variant, that variant was filtered. All variants were then annotated with VEP v102 and assigned to a gene-based on the primary MANE select v0.97 transcript^71^ with the most severe consequence. Following variant quality control and processing, we then selected 421,064 individuals of primarily European genetic ancestry for further analysis. We next queried quality-controlled variant call files for all variants within *DDX3X* (MANE transcript NM_001356.5) identified as SGE-enriched/SGE- depleted. This query yielded a total of 18 variants across 37 individuals, of which 16 and 2 were SGE-enriched or slow-depleting, respectively. Due to the low overall number of slow-depleting *DDX3X* variants, further analyses were limited to SGE-enriched variants.

To determine the phenotypic consequences of carrying such variants, we queried UK Biobank-provided complete health outcomes data (combines general practitioner records, hospital episode statistics, death records, and self-reported conditions), cancer registry data and a subset of binary and continuous phenotypes deemed likely relevant to *DDX3X* loss or gain-of-function (Supplementary Table 7). We then tested whether carrying an ‘enriched’ *DDX3X* variant led to a significant increase in risk for any of the conditions or phenotypes outlined above. Models were implemented in pythonv3.7 using the ‘statsmodels’ package ^72^ with family set to ‘binomial’ or ‘gaussian’ if the trait was binary or continuous, respectively. All models were corrected for age, age^2^, sex, WES sequencing batch, and the first ten genetic principal components as defined in Bycroft et al.^73^

### Analysis of observed vs expected numbers of variants in UKBB and GnomAD

A triplet-based neutral mutational model^30^ was used to estimate the likelihood of each *DDX3X* SNV arising per generation. Analysis was limited to SNVs occurring within 10bp of an exon-intron boundary to ensure exome data coverage. The sum of mutational probabilities for SNVs in each SGE functional class (SGE-unchanged, fast-depleting, slow-depleting, SGE-enriched) was calculated. Across UKBB and GnomAD there are 520 observations of *DDX3X* SNVs (counting a variant observed in both databases as two observations). To estimate the number of SNVs in each SGE functional class that would be expected if the SNVs arose according to the neutral mutational model, the total number of observations of *DDX3X* SNVs was multiplied by the summed mutational probability for each SGE functional class. Supplementary Table 8 gives the mutational probability for each SNV in *DDX3X* as per the triplet-based neutral mutational model.

### Training and assessing a machine learning classifier for NDD-relevance

A supervised Random Forest classifier, built using scikit-learn in Python^37^ was trained on variant LFC. Default parameters were used for bootstrap sample size, the number of features randomly sampled for each split point, the number of trees and tree depth, as modification of the default did not improve model performance. Variants observed in the GnomAD and UKBB population databases and benign/likely benign variants deposited in ClinVar and DECIPHER were used as ‘true negative’ variants (n=540). ‘True positive’ variants (n=168) were *de novo* pathogenic/likely pathogenic variants curated as described above. Stratified sampling was used to select 80% of these variants to train the model. The class with the highest mean probability estimate across the trees in the forest model was taken as the model-predicted class. Variants categorised with the true positive training variants will be referred to as functionally abnormal, and variants categorised with the true negative training variants will be referred to as functionally normal^74^. Model performance was evaluated on the remaining 20% of variants. True positive (TP) variants were variants predicted as non-functional by the clinical classifier, and present in the test set of *de novo* pathogenic/likely pathogenic variants. True negative (TN) variants were variants predicted as functional, and present in the test set of GnomAD/UKBB/clinical benign variants. GnomAD/UKBB/clinical benign variants called as non-functional were considered false positives (FP), while *de novo* likely/pathogenic variants called as functional were considered false negatives (FN). Model sensitivity and specificity increased by incorporating log fold-change data from multiple timepoints. The best performing model incorporated variant LFC on Day 7, 11 and 15. The addition of variant log fold-change on Day 21 did not further improve the performance of the model (Supplementary Table 9). Supplementary Table 9 gives the average number of true and false positives and negatives, sensitivity, specificity, positive and negative predictive values of ten independent versions of each model, evaluated on the test data.

### Estimation of the proportion of *DDX3X* missense variants pathogenic for DDX3X-related NDD

If all missense variants are pathogenic, then the proportion of missense:nonsense variants observed in a large disease cohort is expected to be equal to that predicted by the germline mutational model. According to the neutral mutational model, we expect 20.5x more missense than nonsense variants. Using the data from over 31,000 NDD families^1^, 72 *de novo* missense and 20 *de novo* nonsense variants were observed. Therefore, we estimate that ∼17.5% of missense variants in *DDX3X* are likely to be pathogenic for NDDs.

### Comparison of SGE clinical classifier to *in silico* variant effect prediction programmes

Thresholds were chosen according to the minimum thresholds required for utilisation of *in silico* variant effect prediction scores as “supporting” evidence of variant effect identified by Pejaver *et al.*^38^ as follows: SIFT < 0.0001: test-positive, SIFT >= 0.001: test-negative^75^. PolyPhen2 >0.978: test-positive, PolyPhen2 <=0.978: test-negative. REVEL > 0.773: test-positive, REVEL <=0.773: test-negative, CADD > 25.3: test-positive, CADD <= 25.3: test-negative. Default developer-recommended thresholds were used for *in silico* variant effect prediction tools as follows: SIFT < 0.05: test-positive, SIFT >= 0.05: test-negative. PolyPhen2 >0.902: test-positive, PolyPhen2 <=0.902: test-negative. REVEL > 0.5: test-positive, REVEL <=0.5: test-negative, CADD > 20: test-positive, CADD <= 20: test-negative. Results are shown in Supplementary Table 10.

### Modelling the impact of SGE data on clinical variant interpretation

The Clinical Genome Resource (ClinGen) Sequence Variant Interpretation (SVI) Working Group has published guidelines for the incorporation of functional assays into clinical variant interpretation^6^. We evaluated the performance of our SGE data according to these guidelines and established that a variant with an SGE clinical classification of non-functional would merit application of the PS3 code during clinical variant interpretation (strong evidence that a variant is pathogenic), while a variant with an SGE clinical-classification of functional would merit application of the BS3 code during clinical variant interpretation (strong evidence that a variant is benign). See Supplementary Table 11, adapted from Supplementary Table 1 of Brnich *et al*. To assess the impact of including SGE data in clinical variant interpretation we performed *in silico* interpretation of all possible synonymous, missense, splice A/D and nonsense variants, and those noncoding and frameshift variants included in the SGE HDR template library in the context of each variant arising in a female proband with moderate intellectual disability. The exercise was performed twice, considering each variant as having arisen *de novo* (Supplementary Table 12) or where the inheritance of that variant is unknown (Supplementary Table 13). The codes were assigned as described in Supplementary Note 1. The quantitative scoring framework outlined by Tavtigian *et al.* ^39^ was used to sum across criteria and classify a variant as benign, likely benign, of uncertain significance, likely pathogenic or pathogenic.

### Genotype-Phenotype correlation

Vineland Adaptive Behaviour Composite scores from three studies were identified ^20, 35, 36^, Supplementary Table 14. Where necessary, genomic coordinates were obtained for published patient variants using the Mutalyzer position converter tool (https://mutalyzer.nl/position-converter). If the variant was reported as a cDNA nucleotide change without specifying the ID of the relevant transcript^20^, transcript NM_001356.4 was used to map the variant to genomic coordinates. Genomic coordinates were checked against reported amino acid changes. The Kolmogrov-Smirnov test (KS-test) indicated that Vineland composite scores were normally distributed, therefore two-tailed T-tests were performed. As these studies used a mixture of the Vineland 2nd and 3rd editions, ANCOVA analysis incorporating patient age and Vineland edition as covariates was performed. No association between the Vineland Adaptive Behaviour Composite score and whether a *DDX3X* variant was SGE-fast or SGE-slow depleting was identified. The following python packages were used for statistical analysis: two-tailed T-test and KS-tests were performed in SciPy^76^; ANCOVA was performed using pingouin^77^. In order to investigate broader phenotypes, a composite score was devised for the Lennox *et al.* cohort, encompassing brain MRI findings, neurological phenotype, cardiac findings, precocious puberty, the experience of seizures and behavioural assessment according to the scoring metric in Supplementary Table 15. Where data was partially missing (for example, if a patient had not undergone an MRI), the cohort’s median value for that aspect of the score was assigned to that individual. All phenotypic scores for the Lennox *et al.* cohort are available in Supplementary Table 16. Milestone data for unaffected children were drawn from the following sources: Age of walking^78^; Age of talking: ages of healthy English-speaking children with up to 5 words extracted from http://wordbank.stanford.edu/ ^79^(a ‘word’ was considered to be any sound used with meaning, such as ‘baa’ for sheep). Supplementary Table 17 and 18 give the age of walking and talking for DDD probands and unaffected children, respectively.

### DDX3X variants in cancer

*DDX3X* missense variants in 90,279 independent cancer samples were obtained from cBioPortal^51, 52^. The Intogen resource was used to identify whether *DDX3X* had been identified as a driver gene (https://www.intogen.org/search?gene=DDX3X)^53^. *DDX3X* was considered a driver gene if a minimum of two methods had identified it as such. *DDX3X* was considered to be not a driver if no methods had identified it as such. Thus *DDX3X* was identified as a driver gene in medulloblastoma, pilocytic astrocytoma, chronic lymphoblastic lymphoma, and lymphoma. The proportion of fast and slow depleting missense variants in *DDX3X*-driver and *DDX3X*-non-driver cancers were compared.

### dNdSCV analysis

dNdScv estimates the ratio of non-synonymous to synonymous variation (dN/dS ratio) to identify likely driver genes where there is an excess of non-synonymous variation. For a given cancer sample, dNdScv estimates the background mutation rate of each gene by incorporating the gene-specific synonymous mutation rate with the variation in mutation rates across genes and the epigenomic context, while controlling for genic sequence composition and mutational signatures. dNdScv can also be utilised to estimate the proportion of cancer-associated missense variants likely to be driver variants.^54^ As *DDX3X* has been reported to act as a male-specific driver gene in some cancers^49, 80^, dNdScv was first run on male and female samples from each tumour type separately. Where non-synonymous substitutions in *DDX3X* occur at a significantly higher (p<0.05) than the expected rate, we considered *DDX3X* to be a driver gene (Supplementary Table 19). *DDX3X* was identified as a driver gene in medulloblastoma and lymphoid tumours in both sexes, melanoma of the skin in males only, and uterine corpus endometrial carcinoma and cervical squamous cell carcinoma/endocervical adenocarcinoma in females, consistent with the previous literature.^15–17, 43–49^ *DDX3X*-driver and *DDX3X*-non-driver datasets were pooled and dNdScv re-run to estimate the proportion of missense variants likely to be driver variants. It is likely that there are further tumours where *DDX3X* acts as a male-specific driver gene (p-value for *DDX3X* substitutions = 0.08, for indels = 0.001 in pooled male “*DDX3X*-non-driver” tumour samples) Supplementary Table 20.

## Supplementary Files and Tables

### Supplementary Note

Supp_File1_TWIST_oligo_pool_sg.txt

Supp_File2_TWIST_oligo_pool_sg2.txt

Supp_File3_pmin-U6-ccdb-hPGK-puro.gb

Supp_File4_O00571_UniRef90_blast.filt

Supp_File5_O00571.conserv

Supp_Table1_TWIST_oligo_amplification.xlsx

Supp_Table2_sgRNA.xlsx

Supp_Table3_HDR plasmid library cloning.xlsx

Supp_Table4_Sequencing_library_prep.xlsx

Supp_Table5.txt

Supp_Table6_All_clinical_variants.txt

Supp_Table7_PHEWAS_enriched_ICD10.tsv

Supp_Table8_DDX3X_all_snvs_mutation_probs.txt

Supp_Table9_Clinical_classifier_outcomes.txt

Supp_Table10_Sensitivity_specificity_default_thresholds.txt

Supp_Table11_Brnich_Odds_Path.xlsx Supp_Table12_Modelling_SGE_data_on_insilico_denovo_var_clinical_classification.txt Supp_Table13_Modelling_SGE_data_on_insilico_inh_unknown_var_clinical_classification.tx t

Supp_Table14_NgCordell_Tang_Lennox_VAB.txt

Supp_Table15_Scoring_for_Lennox_composite_phenotypic_score.xlsx Supp_Table16_LENNOX_composite_phenotypic_score.txt

Supp_Table17_DDD_DDX3X_pt_phenotypes_talking.txt

Supp_Table18_DDD_DDX3X_pt_phenotypes_walking.txt

Supp_Table19_dNdScv_outputs_cancers_split_by_sex.txt

Supp_Table20_dNdScv_outputs_pooled_cancers.txt

---

**Extended Data 1:**
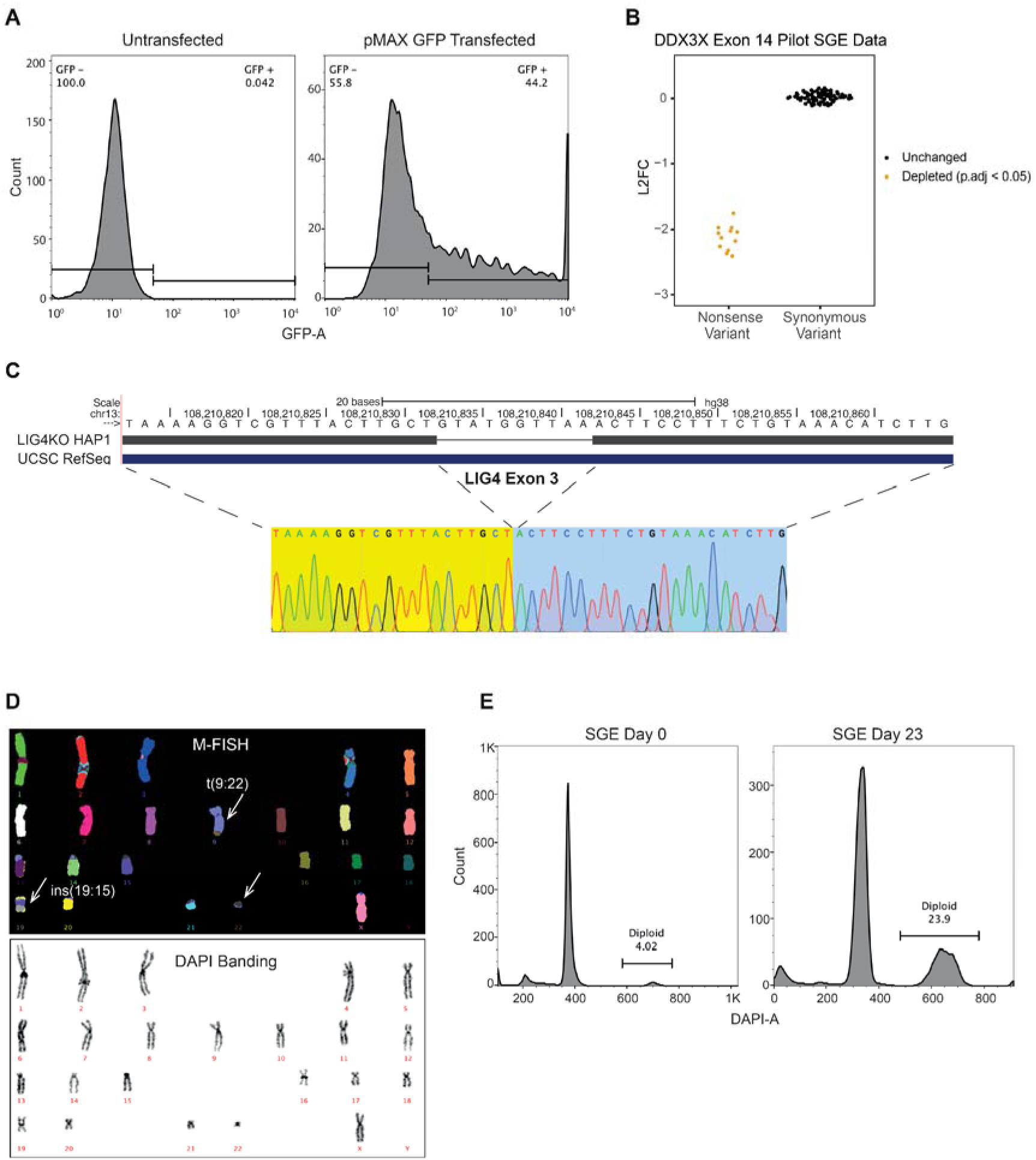
Optimisation of Saturation Genome Editing. A) Estimation of co-transfection efficiency of the SGE protocol. pMin-U6-DDX3XsgRNA-hPGK-puro and pMAX GFP plasmid were used for the co-transfection. Representative FACS plot showing 44% co-transfection efficiency. Only the viable singlet cells were used for the analysis. B) Pilot SGE study of *DDX3X* exon 14. Day 11 vs Day 4 Log2 fold-change of nonsense and synonymous variants. C/D) HAP1 cell bank validation: C) Sanger sequencing shows 10bp deletion within exon 3 of the *LIG4* gene. D) Representative karyotyping by M-FISH and DAPI banding showing haploid cells with the expected t(9;22) and ins(15;19) karyotype. E) On Day 0 of a representative SGE experiment, the percentage of diploid cells was 4.07%. On Day 23 of a representative SGE experiment, the percentage of diploid cells was 23.9%. Only the singlet cells were used for the analysis.

**Extended Data 2:**
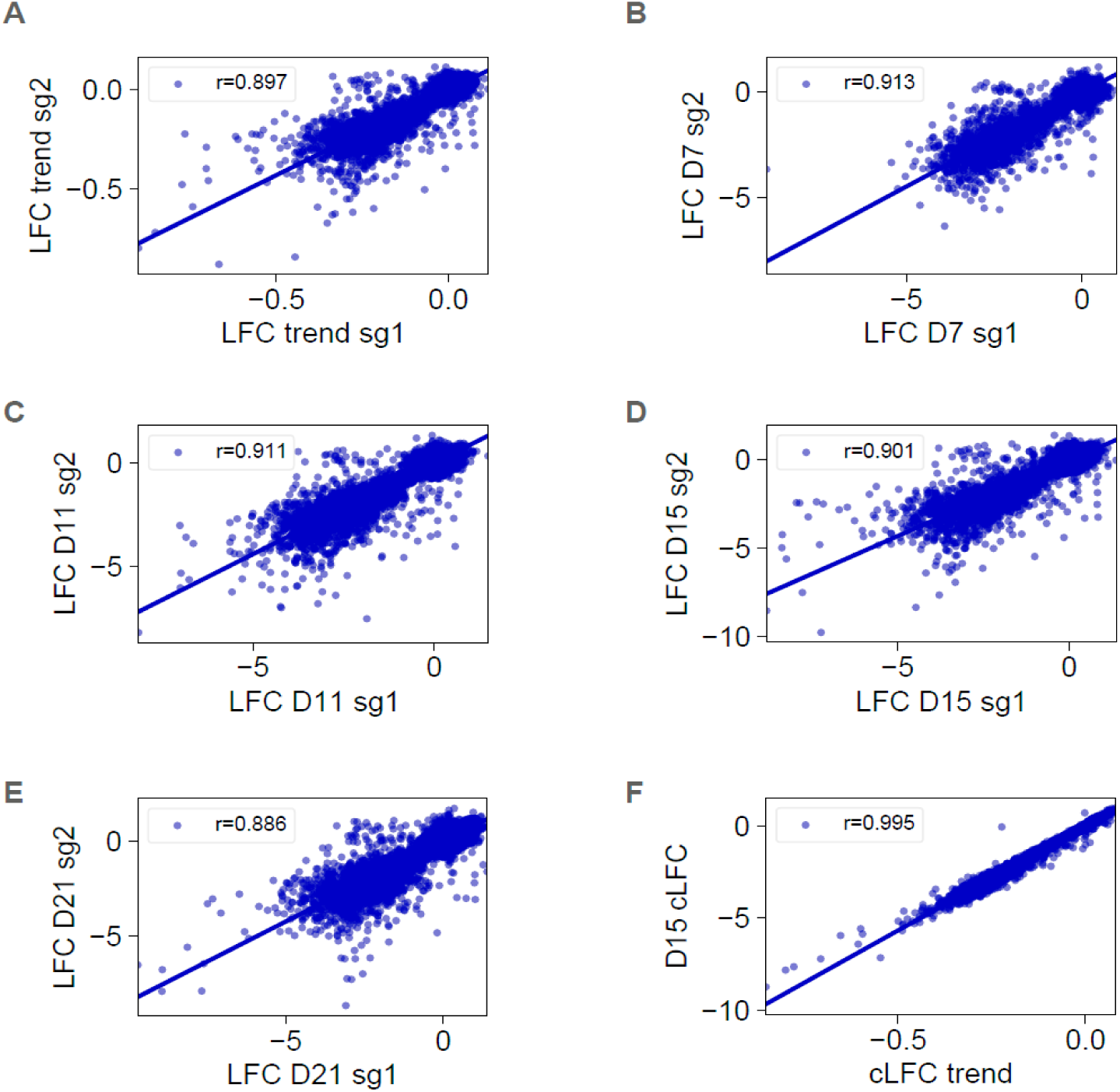
Correlation between sgRNAs and between measures of variant abundance. A) Correlation between LFC-trend for sg1 and sg2; Pearson r=0.897. B) Correlation between Day 7 LFC for sg1 and sg2; Pearson r=0.913. C) Correlation between Day 11 LFC for sg1 and sg2; Pearson r=0.911. D) Correlation between Day 15 LFC for sg1 and sg2; Pearson r=0.901. E) Correlation between Day 21 LFC for sg1 and sg2; Pearson r=0.886. F) Correlation between Day 15 cLFC and cLFC trend; Pearson r=0.995.

**Extended Data 3:**
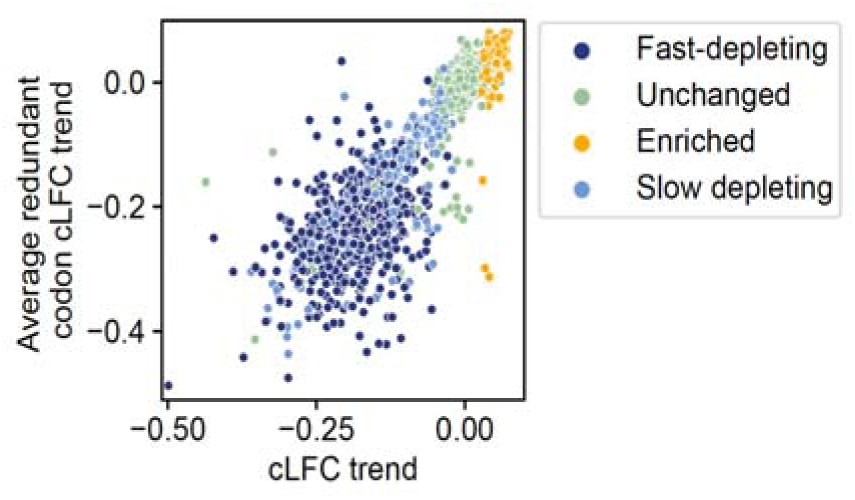
Correlation between cLFC trend and average redundant codon cLFC trend.

**Extended Data 4:**
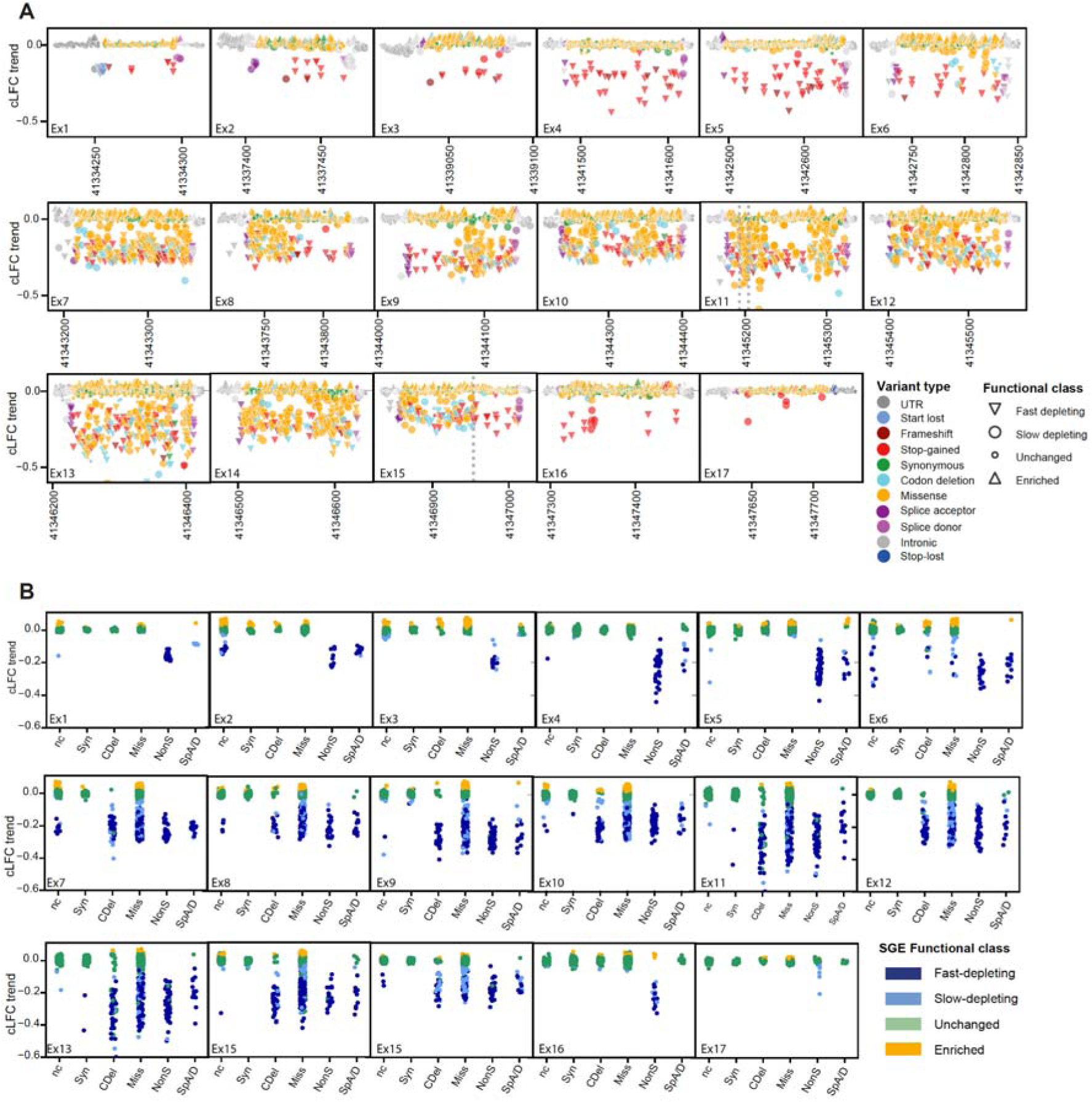
cLFC trend of all *DDX3X* variants. A) y-axis: cLFC-trend; x-axis: Hg38 position, grouped by exon. Variants coloured by variant type, shape indicates SGE functional class. B) cLFC-trend of non-coding (nc), synonymous (Syn), in-frame codon-deletion (Cdel), missense (Miss), nonsense (NonS) and canonical splice acceptor/donor variants (SpA/D), grouped by exon and coloured by SGE functional class.

## Supplementary Note

### *In silico* modelling of clinical variant interpretation and the impact of SGE functional data incorporation

Codes were assigned as follows, according to the ClinGen SVI Guidelines: BS1 criterion:

- Moderate if variant not observed in either UKBB or GnomAD datasets.
- Strong if allele frequency > 3.25*10^-7 in any of the major population groups of GnomAD or UKBB. The threshold was estimated as 3.25*10^-7 using http://cardiodb.org/allelefrequencyapp/ ^1^ as follows:
- Prevalence of DDX3X-related DD: 1/27700, calculated as 1% (population prevalence of ID) x prevalence of *DDX3X de novo* in the Kaplanis *et al.* (112/31058)^2^, which is consistent with previous estimates.
- Allelic heterogeneity: estimated as 0.09. In Kaplanis *et al.*, the most common variants were observed 5 times, with a total of 112 different *de novo DDX3X* variants, 0.09 is the upper bound of the CI^2^.
- Genetic heterogeneity: 1 (only *DDX3X* has been reported to cause *DDX3X*-related neurodevelopmental syndrome).
- Penetrance: 0.5 (likely to be an under-estimate).

PP3/BP4 criteria:

- PP3 - supporting if SpliceAI Delta score > 0.7 or REVEL score > 0.7, or if REVEL score not available if CADD PHRED score > 25
- BP4 - supporting if SpliceAI Delta score < 0.2 and if REVEL score < 0.2, or if REVEL score not available, if CADD PHRED < 10.

PS2 criterion:

- Supporting if the variant is not *de novo* but this variant has previously been reported as *de novo* and pathogenic or likely pathogenic
- Moderate if the variant is *de novo* and this variant has previously been reported as *de novo* and pathogenic or likely pathogenic

PVS1 criterion

Nonsense and frameshift variants

- Very strong if the variant is predicted to undergo NMD
- Strong if the variant is not predicted to undergo NMD, occurs distal to the DDX3X helicase domains, and results in truncation of > 10% of the protein. (LOFs are not frequent in the general population, including in the last 10% of the protein)
- Moderate if the variant is not predicted to undergo NMD, occurs distal to the DDX3X helicase domains, and results in truncation of < 10% of the protein.

Canonical splice acceptor/donor variants (PP3 was not applied to avoid double-counting of information):

- Very strong if the exon is out-of-frame so exon skipping would be predicted to result in a frame-shift, and the variant is predicted to undergo NMD.
- Strong if the exon is out-of-frame, the variant is not predicted to undergo NMD, and the resultant frame-shift caused by exon skipping would result in truncation of > 10% of the protein.
- Moderate if the exon is out-of-frame, the variant is not predicted to undergo NMD, and the resultant frame-shift caused by exon skipping would result in truncation of < 10% of the protein.
- Moderate if the exon is in-frame, the variant occurs outside of the regions encoding the DDX3X helicase domains. (All DDX3X exons are < 10% of the length of the coding length).

The following variants were predicted to escape NMD:

- occurring within the first 200bp of the coding sequence of *DDX3X*
- occurring distal to 55bp 5’ of the last coding splice junction (between exons 16 and 17) of *DDX3X*.

Missense variant-specific criteria:

- PP2 - supporting for all variants, as missense variants are frequently pathogenic in DDX3X-related neurodevelopmental syndrome and DDX3X is known to be missense-constrained.
- PS1 - strong, if the variant resulted in the same amino-acid change as a previously reported *de novo* pathogenic or likely pathogenic variant.
- PM5 - moderate, if the variant resulted in a different amino acid at a residue with a previously reported *de novo* pathogenic or likely pathogenic variant.
- PP5 - supporting if a non-*de novo* variant had been reported as pathogenic or likely pathogenic at this amino acid residue, regardless of the match of the amino acid change.

